# Assessment of effective mitigation and prediction of the spread of SARS-CoV-2 in Germany using demographic information and spatial resolution

**DOI:** 10.1101/2020.12.18.20248509

**Authors:** Martin J. Kühn, Daniel Abele, Tanmay Mitra, Wadim Koslow, Majid Abedi, Kathrin Rack, Martin Siggel, Sahamoddin Khailaie, Margrit Klitz, Sebastian Binder, Luca Spataro, Jonas Gilg, Jan Kleinert, Matthias Häberle, Lena Plötzke, Christoph D. Spinner, Melanie Stecher, Xiao Xiang Zhu, Achim Basermann, Michael Meyer-Hermann

**Affiliations:** Institute for Software Technology, German Aerospace Center, Cologne, Germany; Department of Systems Immunology and Braunschweig Integrated Centre of Systems Biology (BRICS), Helmholtz Centre for Infection Research, Braunschweig, Germany; Earth Observation Center, Department EO Data Science, German Aerospace Center, Weßling, Germany; Technical University of Munich, School of Medicine, University Hospital rechts der Isar, Department of Internal Medicine II, Munich, Germany; University Hospital of Cologne, Department I for Internal Medicine, University of Cologne, Cologne, Germany; German Center for Infection Research (DZIF), Cologne, Germany

**Keywords:** SARS-CoV-2, Covid-19, Coronavirus disease, Mitigation, Non-pharmaceutical interventions, Forecast

## Abstract

Non-pharmaceutical interventions (NPIs) are important to mitigate the spread of infectious diseases as long as no vaccination or outstanding medical treatments are available. We assess the effectiveness of the sets of non-pharmaceutical interventions that were in place during the course of the Coron-avirus disease 2019 (Covid-19) pandemic in Germany. Our results are based on hybrid models, combining SIR-type models on local scales with spatial resolution. In order to account for the age-dependence of the severe acute respiratory syndrome coronavirus 2 (SARS-CoV-2), we include realistic pre-pandemic and recently recorded contact patterns between age groups. The implementation of non-pharmaceutical interventions will occur on changed contact patterns, improved isolation, or reduced infectiousness when, e.g., wearing masks. In order to account for spatial heterogeneity, we use a graph approach and we include high-quality information on commuting activities combined with traveling information from social networks. The remaining uncertainty will be accounted for by a large number of randomized simulation runs. Based on the derived factors for the effectiveness of different non-pharmaceutical interventions over the past months, we provide different forecast scenarios for the upcoming time.

**Mathematics Subject Classification (2010)** 00A72 *·* 65L05 *·* 68U20

## 1 Introduction

With more than 1.6 million reported deaths [46], the coronavirus disease 2019 (Covid-19) remains one of the most pressing issues for the whole globe. Already in October, WHO officials estimated that 10 % of the world’s population had been infected [30] and vaccination of the population will still take a considerable amount of time. Since exposing people is highly unethical [91], the only interim solution is to mitigate the spread of the disease by the application of non-pharmaceutical interventions.

The assessment of non-pharmaceutical interventions and prediction by simulation has to be based on reliable models; cf. [5, 6, 23, 29, 35, 53, 63, 82, 13] for the spread of SARS-CoV-2 (severe acute respiratory syndrome coronavirus 2) and other infectious diseases. Only then, the most effective interventions can be determined as a basis for informed political decisions.

The aim of our study is to assess non-pharmaceutical interventions and to provide a reliable forecast of the Covid-19 pandemic in Germany based on four principles. First, we account for the age-dependence of SARS-CoV-2 [67, 59, 87]. Second, we include realistic contact patterns between different age groups [62, 37, 68, 45, 47]. Third, we include high-quality, spatially resolved information on commuting activities [10, 12] combined with traveling information based on the social network Twitter. Fourth, we combine all information and account for the remaining uncertainty by Monte-Carlo Ensemble runs. To our knowledge, such an in-depth study is not accounted for in the literature so far.

The remainder of this paper is structured as follows. In Section 2, we present our mathematical model and its numerical solution approach. In Section 3, we present the social and non-pharmaceutical parameters used in our model, and in Section 4, we discuss the epidemiological parameters obtained by extensive analyses. Our results are presented and discussed in Section 5. We conclude this paper with Section 6.

## 2 Model and solver

There are various models to forecast the spread of infectious diseases across a country or community. Besides the well known SIR-type ODE models [52, 3], there are integro-differential models [52, 51], Bayesian Monte Carlo approaches [82], or agent-based models [55, 9]. While SIR-type models are praised for their simplicity and understandability, they lack for a good representation of spatial heterogeneity.

To combine the advantages of a SIR model without loss of spatial resolution, we use multiple SIR-type models on a fine local scale and connect the compartments and age groups by graphs that represent traveling; cf. [2] among others. These SIR-type models can be easily exchanged by agent-based models due to the generic implementation of our graphs. This is integrated as part of our *high performance modular epidemics simulation* software *MEMILIO* that is continuously under development [1].

### 2.1 Age-resolved SIR-type model

The base of our SIR-type model can be found in the first version of [53]. Our model consists of the compartments *Susceptible* (S), healthy individuals without immune memory of SARS-CoV-2; *Exposed* (E), who carry the virus but are not yet infectious to others; *Carrier* (C), who carry the virus and are infectious to others but do not yet show symptoms (they may be pre- or asymptomatic); *Infected* (I), who carry the virus, are infectious and show symptoms; *Hospitalized* (H), who experience a severe development of the disease; *In Intensive Care Unit* (U); *Dead* (D); and *Recovered* (R), who cannot be infected again. To resolve age-specific disease parameters, we divide the totality of people *N* into *n* different age groups. We then have

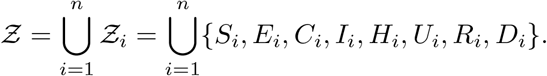

For each age group *i* = 1, …, *n*, the transmission risk is denoted by *ρ*_*i*_ and the proportion of infected people not isolated or quarantined is denoted by 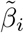; see Tables 1 and 2 for details. Infection results from contact with people from different age groups. We introduce the nonsymmetric contact frequency matrix

**Table 1.** Description of parameters and main resources for their derivation; see Table 2 for the age-dependent ranges of the parameters.

| Param. | Description | Reference | Resources |
| --- | --- | --- | --- |
| $\rho^{(0)}$ | $\rho_i$ age-dependent transmission risk | Eq. (14),(2),(3) | [41,80,11,57,93,28,36] |
| $k$ | seasonality parameter | Eq. (15), (14) | [81,88,69,65] |
| $\tilde{\beta}$ | proportion of not isolated or quarantined symptomatic individuals | Eq. (2), (3) | Assumption. |
| $T_E^C$ | period of latent non-infectious stage | Eq. (3), (4) | [53,64,94] |
| $\mu_C^R$ | proportion of mild, asymptomatic cases | Eq. (4), (5), (8) | [48,22,14,28,95] |
| $T_C^R$ | period of asymptomatic stage before recovery | Eq. (4), (8) | [53] |
| $T_C^I$ | period of latent infectious stage | Eq. (4), (5) | [53,64,94] |
| $\mu_I^H$ | proportion of symptomatic cases needing hospitalization | Eq. (5), (6), (8) | [72,87,34] |
| $T_I^H$ | period of mild symptoms for individuals requiring hospitalization later on | Eq. (5), (6), Fig. 16 | [32,54,42] |
| $T_I^R$ | period of mild symptoms for individuals not requiring hospitalization later on | Eq. (5), (8) | [90,53] |
| $\mu_H^U$ | proportion of hospitalized individuals getting ICU treatment | Eq. (6), (7), (8) | [89,50,34] |
| $T_H^U$ | period of hospitalization before ICU treatment (of critical cases) | Eq. (6), (7), Fig. 17 | [32,54] |
| $T_H^R$ | period of hospitalization before recovery (of non-critical cases) | Eq. (6), (8), Fig. 18 | [53] |
| $\mu_U^D$ | proportion of individuals in ICU care that die | Eq. (7), (8), (9), Fig. 6 | [50,4] |
| $T_U^R$ | period of ICU treatment before recovery | Eq. (7), (8), Fig. 19 | [53,92,54] |
| $T_U^D$ | period of ICU treatment before death | Eq. (7), (9), Fig. 20 | [54,53] |

**Table 2.**
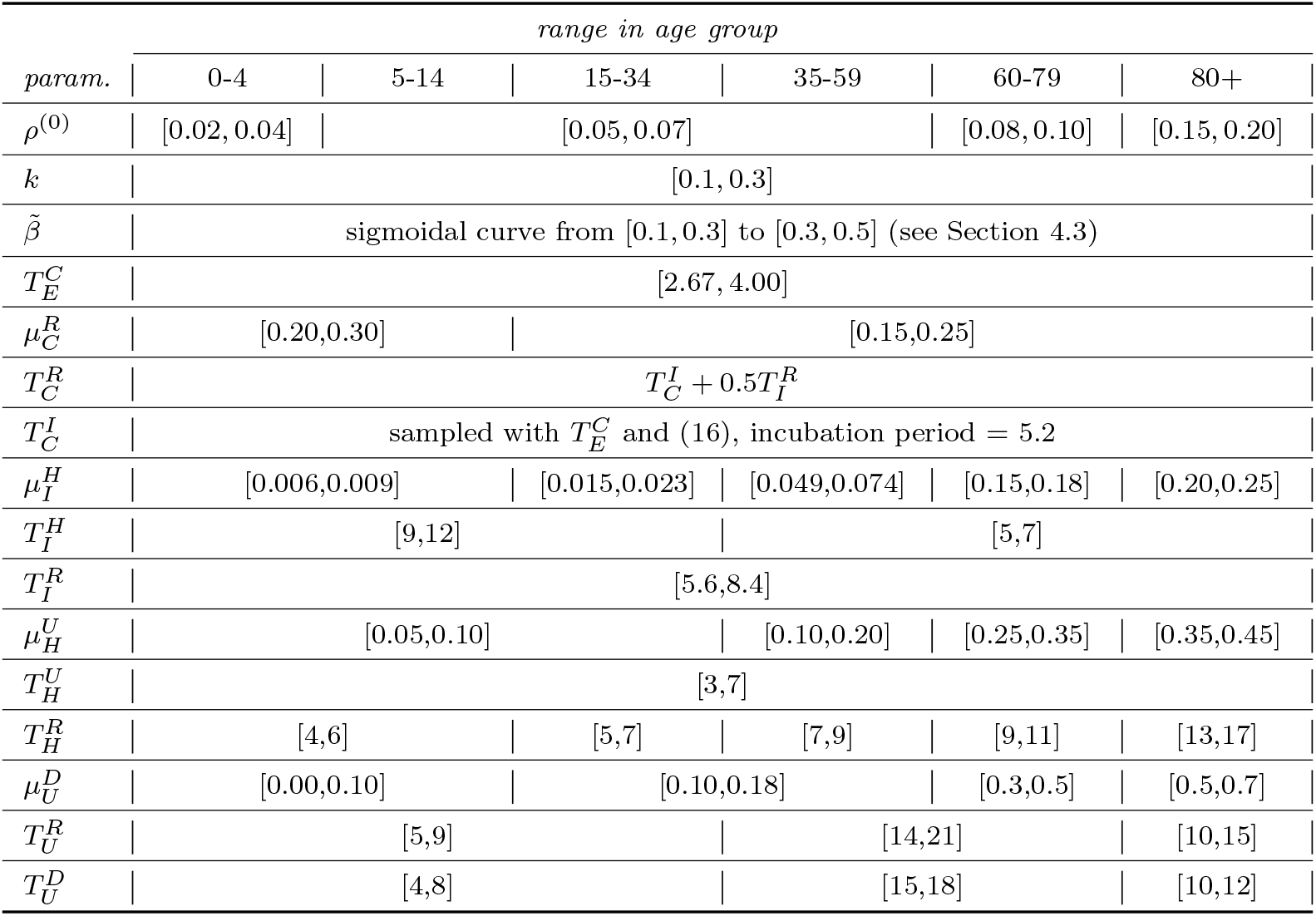
Summary of the age-dependency of parameters and their ranges; see Table 1 for the parameters’ description and the references from which they are derived.

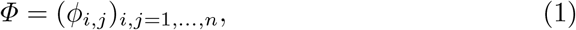

where *ϕ*_*i,j*_ represents the (mean) daily contacts of a person of age group *i* with people from age group *j*. Since contact patterns are assumed to be nonsymmetric – think of contact rates of a group of school children with their teacher and vice versa – this matrix will also be nonsymmetric.

The naming convention for the remaining parameters can be understood as follows: We use the variables 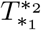 for the time spent in state *∗*_1_ *∈𝒵*_*i*_ before moving to state _2_ *∈𝒵*_*i*_. For example, 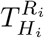 represents the time an individual in age group *i* = 1, …, *n* spent in the hospital before returning home due to recovery from the disease. Accordingly, 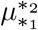 represents the probability of a patient to *transit* to state *∗*_2_ when that patient is currently in state *∗*_1._

The model, as expressed in Figure 1, is

**Fig. 1.**
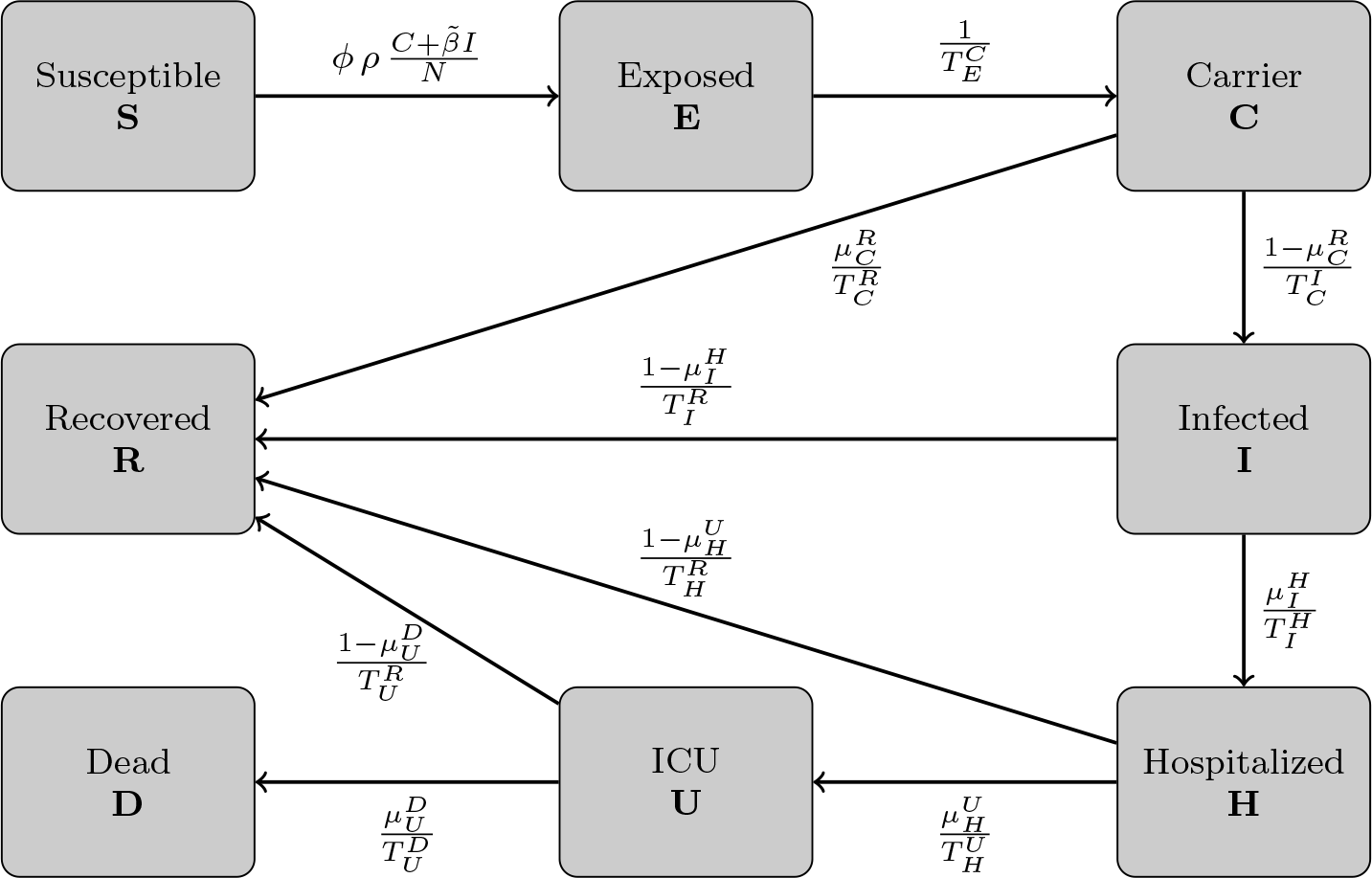
SIR-type model based the on first version of [53]. We omit the age-dependence index *i* for clarity; see Table 1 and 2 for a description of the parameters.

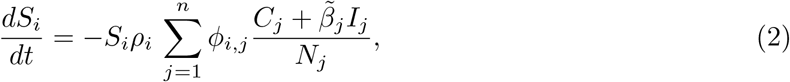

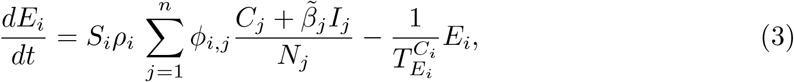

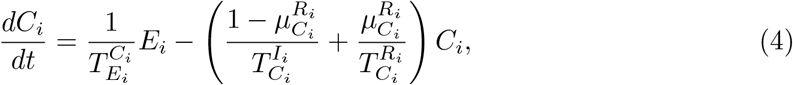

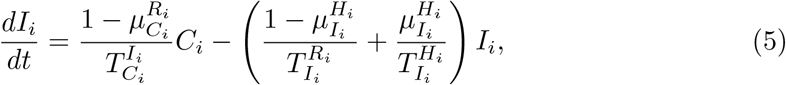

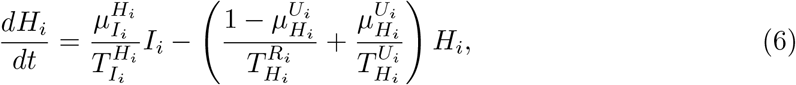

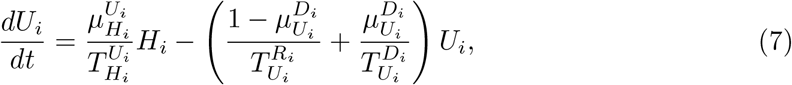

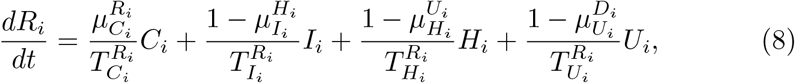

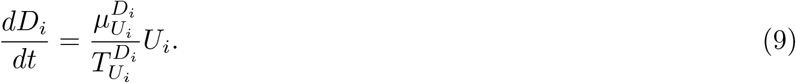

The equations (2)–(9) represent the transition of people from one state to another. Note that people in an age group *𝒵*_*i*_ cannot *transit* to another group 𝒵_*j*_ for *i* ≠ *j*. In Section 4, we will discuss in detail which of the epidemiological parameters we assume to be age-dependent and how this is included in our model. These findings are summarized in Tables 1 and 2.

### 2.2 Spatial resolution

While SIR-type models are straightforward to apply and interpret, they lack the possibility of modeling local effects or spatial heterogeneity. In order to avoid averaging over important effects such as infection clusters, we assign one particular age-resolved model to each county. We represent each county by a node of a (directed) graph. The edges of the graph represent the connections between the different counties and are weighted with the number of people commuting daily and traveling on average. The edges do not only hold single values (weights) for how many people daily commute between different counties but also coefficients to determine the proportion of people of different age groups and compartments that commute or travel. Doing so, we can restrict travel activities to healthy or only mildly infected individuals.

Let *n*_*C*_ be the number of counties (nodes of the graph). Then, for two nodes *a*_*k*_ and *a*_*l*_, 1 *≤ k, l ≤ n*_*C*_, the weight *w*_*k,l*_ on edge *e*_*k,l*_ represents the proportion of people going daily from *a*_*k*_ to *a*_*l*_; see Figure 2 (left).

**Fig. 2.**
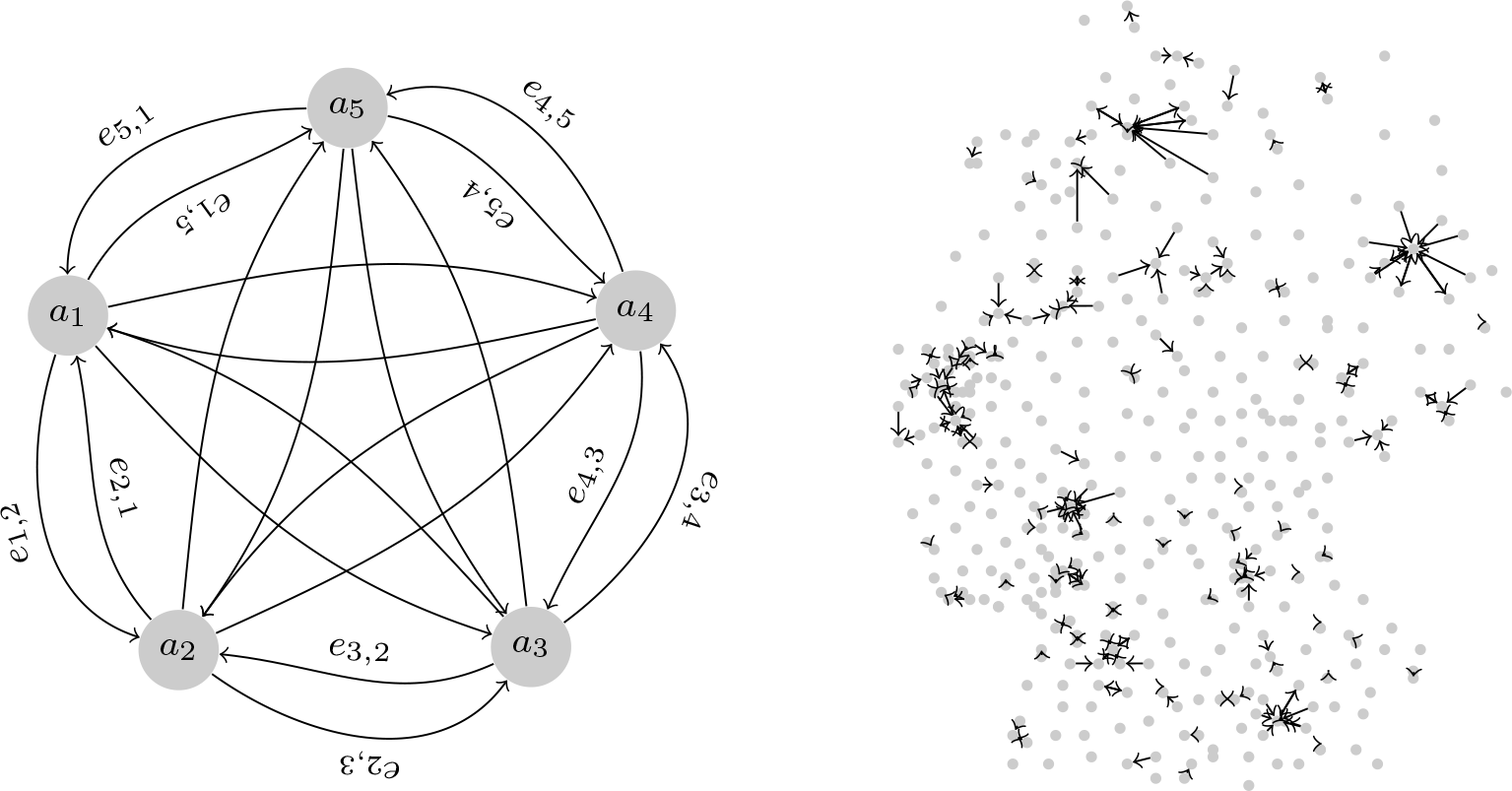
**Left:** Graph model with five nodes. Each node is assigned one particular age-resolved SIR-type model. For each county, represented by node *a*_*k*_, there is an edge *e*_*k,l*_ to node *a*_*l*_. Edge descriptions not shown in the interior. **Right:** Center points of all German counties. Edges only shown where more than 10 000 workers commute on a daily basis.

### 2.3 Numerical solver

Common numerical solvers for the system of nonlinear ordinary differential equations (2)-(9) are semi-implicit or adaptive explicit. While the former allow for larger time steps, the latter allow adaptive time steps to prevent large numerical errors. We have implemented an adaptive Runge-Kutta-Fehlberg45 (RKF45) method [33] that uses methods of 4th and 5th order and solves the equations without excessively small time steps.

The numerical procedure becomes more challenging when we also resolve the equations spatially. For this, we define a commuter as a person who travels from county *a*_*k*_ to *a*_*l*_, 1 ≤ *k, l ≤n*_*C*_ and back again within one day (whether it is work or free time related). Given start values from day *t*, we advance our adaptive RKF45 solver for 0.5 days. Next, we allow people to commute or travel. Their amount is defined by the commuter rate between two counties, namely the weights *w*_*k,l*_ introduced in Section 2.2 and further specified in Section 3.1. Note that commuting also depends on the infection state since hospitalized individuals cannot commute and infected individuals will travel less than healthy ones. For the latter, we assume the same level of isolation or quarantine as on county level. With the updated population, we again advance our adaptive solver for 0.5 days.

Additionally, we conduct an auxiliary step with step size of 0.5 days with an explicit Euler solver where we only consider the in-commuters, using the county’s population as contact population only. This step is executed since, after the high precision scheme from *t*+0.5 to *t*+1, we do not know the updated state of our commuters (e.g., susceptible may have become exposed or carriers have become symptomatic). This is due to the nature of the SIR model (2)-(9) that does not keep track of individuals. Still, the commuters have to go back to their home county, and we need to know their most likely infection state. We use the results from the explicit Euler step, to quantify the proportion of individuals of the different compartments that return. With this estimation, we start the returning process. These considerations are summarized in Figure 3.

**Fig. 3.**
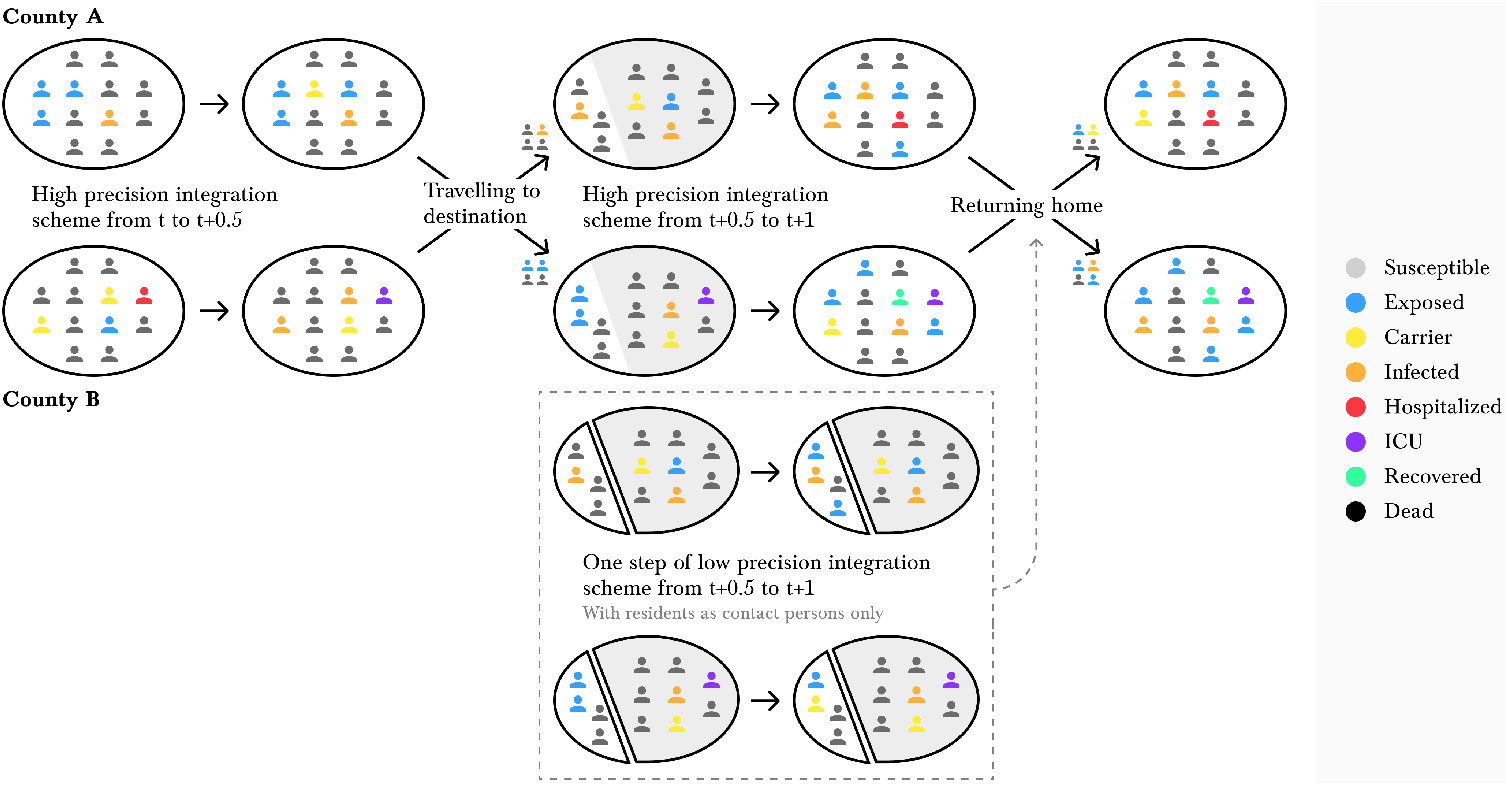
Illustration of our numerical scheme for spatial heterogeneity. The individuals represent the compartments in our SIR model (2)-(9). First, we evolve County A and County B separately, resulting in updated compartments in both counties. Then, a proportion of the county’s population commutes. We solve the SIR model for another half day including the new in-commuters and excluding the out-commuters. Additionally, we conduct one step of an auxiliary low order scheme (with residents as contact persons only) to estimate the updated states of the in-commuters (shown in the dashed box). We then allow the commuters to return home.

## 3 Social and non-pharmaceutical parameters

The spread of SARS-CoV-2 depends on many parameters. While some of these parameters are inherent to the virus, others depend on social contact patterns and non-pharmaceutical interventions introduced by decision makers. In this section, we will focus on non-pharmaceutical interventions and their influence on contact patterns and commuter rates in our model.

### 3.1 Inter-county travel

Let us first continue with the spatial resolution of our model and focus on how we specify the rate of work or leisure commuters *w*_*k,l*_ between different counties *a*_*k*_ and *a*_*l*_. To estimate *w*_*k,l*_ on the edge *e*_*k,l*_ that connects both counties, we use statistics from the German Federal Employment Agency [10] complemented with geo-referenced Twitter data.

From [10], we have high quality information on work commuter activities. For each county, we have the number of incoming commuters from each other county. However, it is not specified if commuting takes place on a daily basis or if workers work partially from home. From the raw data, we assume that any commuting activity with linear distance of 100-200 km only happens twice a week and for a distance of over 200 km, we assume that it happens only once a week. That means, we scale the corresponding off-diagonal matrix entries by 0.5 and 0.2, respectively. After this smoothing procedure, we remain with about 11.8 million daily commuting activities. These values are then divided by the approximate population size in working age. This results in the work commuting population *c*_*k,l*_, 1≤ *k, l ≤ n*_*C*_ which is only taken from age groups and compartments that commute for work.

Motivated by [49, 61], we also analyzed a totality of about 2.8 million georeferenced tweets obtained from the Twitter API [85] between January 1, 2018 and January 31, 2020. Since [10] is biased towards the working population, the geo-referenced tweets act as a “counterpart” with a bias towards the younger population, student commuters, and leisure traveling individuals. From the totality of tweets, we first removed all tweets from users with more than 1000 tweets in the considered period to waive statistical anomalies introduced by bots or hyperactive users. For each pair of consecutive user tweets in the considered period, we then count one travel action from *a*_*k*_ to *a*_*l*_ if the first tweet happened from *a*_*k*_ and the second happened from *a*_*l*_, *k* ≠ *l*. In order to further reduce the influence of the most active users, we remove the first 300 users with the most travels. Then, there remain about 36 000 active users from which 11 000 had five or more travel activities in the last two years. In total, we have about 235 700 travel activities. In 95 % of the nontrivial matrix entries, one unique user adds only two activities on average.

Although different in scale, the remaining tweets show similar travel patterns as obtained from [10]; see Figures 21-23 for a comparison. Note, however, that a certain amount of deviation is to be expected since both sources are biased towards either the working or the student population. In twitter mobility, for instance, we observe relatively strong connections to bigger cities like Hamburg, Hannover, Cologne, or Stuttgart.

Assuming that the mobility obtained from Twitter accounts for 20 % of all travel activities (work commuting, student commuting, leisure travel etc.), we scale the twitter matrix accordingly. The resulting values are divided by the population size and the result will be denoted by *t*_*k,l*_, 1 *≤ k, l ≤ n*_*C*_.

The amount of mobility in our model is given be the edge weights *e*_*k,l*_ of the graph. These weights are derived from the matrix *c*_*k,l*_ for the work commuters and from *t*_*k,l*_ for the ‘Twitter’ activities. The weights also depend on the implementation of non-pharmaceutical interventions. In particular, they depend on the NPI related parameters 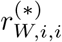 for work and 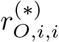 for “other” places related measures that mainly affect free-time activities. Here, * ∈{1, 2} and *i* = 1, …, *n* are the corresponding age groups; see Section 3.3.2 and 3.3.1 and Table 3 for details on these parameters.

**Table 3.** Summary of different non-pharmaceutical interventions and implementations in our simulations; cf. Section 3.4.

| *intervention* | *implementation* | *factor ranges* | *comment* |
| --- | --- | --- | --- |
| working<br>from<br>home | weak<br>intermediate<br>strong | $r_{W,i,j}^{(1)} \in [0.0, 0.1]$<br>$r_{W,i,j}^{(1)} \in [0.2, 0.3]$<br>$r_{W,i,j}^{(1)} \in [0.4, 0.5]$ | |
| partial school<br>closures and<br>remote<br>schooling | none<br>weak<br>intermediate<br>complete | $r_{S,i,j}^{(1)} = 0.0$<br>$r_{S,i,j}^{(1)} = 0.25$<br>$r_{S,i,j}^{(1)} = 0.5$<br>$r_{S,i,j}^{(1)} = 1$ | |
| gathering bans,<br>(partial) closing<br>of bars,<br>restaurants,<br>cinemas etc. | weak<br>rather weak<br>intermediate<br>strong<br>very strong | $r_{O,i,j}^{(1)} \in [0.0, 0.2]$<br>$r_{O,i,j}^{(1)} \in [0.2, 0.4]$<br>$r_{O,i,j}^{(1)} \in [0.4, 0.6]$<br>$r_{O,i,j}^{(1)} \in [0.6, 0.8]$<br>$r_{O,i,j}^{(1)} \in [0.8, 1.0]$ | additional increase of<br>$r_{W,i,j}^{(1)}$ by $[0.0, 0.05]$,<br>$[0.05, 0.10]$, and $[0.1, 0.2]$<br>according to strictness;<br>cf. Section 3.3.2 |
| face masks,<br>distancing,<br>regular<br>ventilation of<br>closed spaces | weak<br>rather weak<br>intermediate<br>strong<br>very strong | $r_{*,i,j}^{(2)} \in [0.0, 0.2]$<br>$r_{*,i,j}^{(2)} \in [0.2, 0.4]$<br>$r_{*,i,j}^{(2)} \in [0.4, 0.6]$<br>$r_{*,i,j}^{(2)} \in [0.6, 0.8]$<br>$r_{*,i,j}^{(2)} \in [0.8, 1.0]$ | $* \in \{H, S, W, O\}$ |

The commuter matrices contain many insignificant coefficients close to zero. These are due to loosely coupled regions where only a very limited number of individuals commute on a daily basis (e.g., 1 or 2). To reduce the computational effort, we eliminate edges *e*_*k,l*_ where *c*_*k,l*_ *<* 4*·*10^*−*5^ and *t*_*k,l*_ *<* 1*·*10^*−*5^. The cutoff values are chosen so only 1 % of information is dropped and that more than 99 % of travels are included. We also paid attention to reflect the above assumption that Twitter data represents 20 % of travels. With this procedure, the number of edges is reduced by approximately 60 % and the computational efficiency is increased significantly.

### 3.2 Contact patterns in Germany

In the following, we focus on the individual counties, interventions and contact patterns. In this section, we derive a baseline pre-pandemic contact matrix 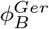 and a minimum contact matrix 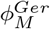 for a simulated strict lockdown in Germany.

As SARS-CoV-2 transmission occurs mainly during human-to-human interaction, reducing contacts can efficiently slow down the spread of the disease; cf. [38, 62, 39, 37, 68] for literature on contacts and the spread of infectious diseases. However, lockdowns which effectively reduce contacts to a minimum should be avoided due to their profound negative impact on many individuals and communities [91]. Therefore, the challenge for many of today’s decision makers is to find the most appropriate and effective interventions for the actual developments.

#### 3.2.1 Pre-pandemic patterns

In order to quantify the potential of transmission reduction by contact pattern changes, good pre-pandemic as well as recent data is needed. From the POLYMOD study [62] and its projections [68], we use realistic contact patterns for Germany split up into the categories “Home”, “School”, “Work”, and “Other”. In [62], contacts are defined as skin-to-skin contact, or where at least three words were exchanged.

For the particular case of school contacts, the mean numbers of contacts recorded in [62] are rather low for Germany; cf. Figure 5 for a comparison of the projection [68] for different European countries. Given the fact of aerosol transmission risk in closed spaces, we suggest to assume slightly higher contact rates for a conservative estimate on the spread of the disease. Further information is offered by the demography-based school contact matrix in [37]. We use the quotient of the maximum eigenvalues between both matrices to scale the contact matrix of [68] which then results in a larger number of school contacts.

**Fig. 5.**
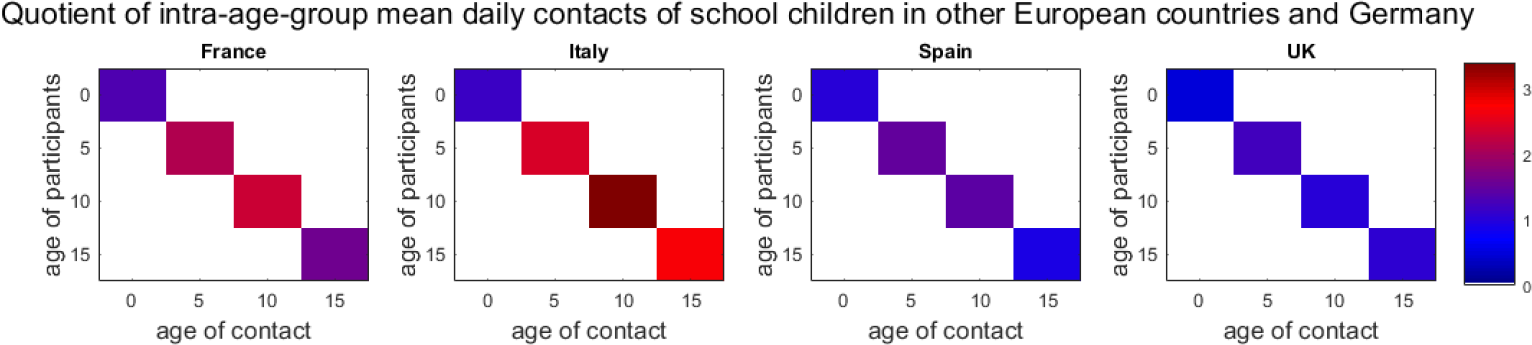
Quotient of recorded, projected [62, 68] intra-age-group mean daily contacts of school children for different European countries and Germany; we, e.g., plot 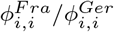.

The combination of baseline contacts for “Home”, “Work”, and “Other” from [62, 68] and for “School” based on the comparison of [68, 37] results in the contact matrix 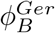; cf. Figure 4 (top).

**Fig. 4.**
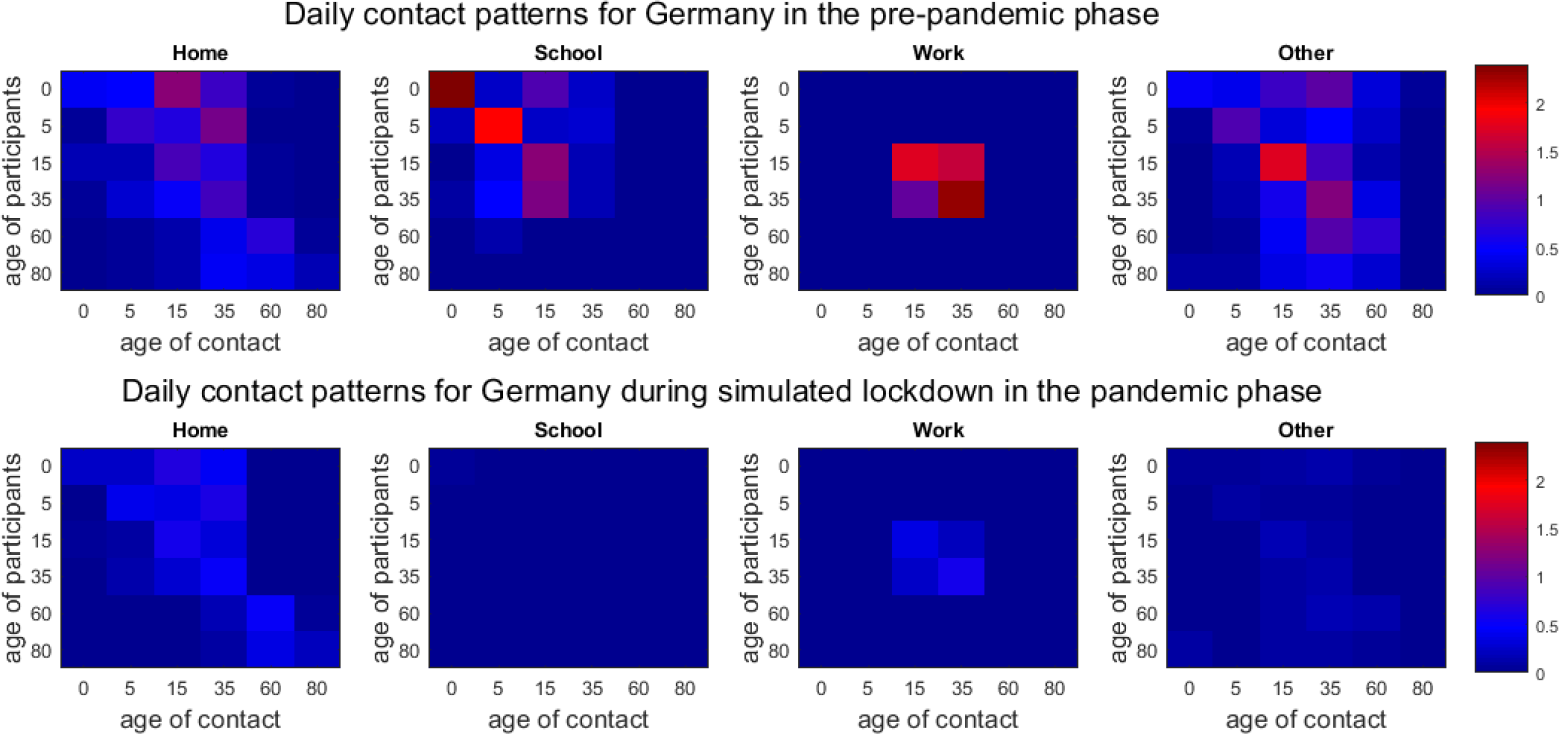
**Top:** Combined pre-pandemic contact patterns 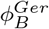 of [68, 37] for Germany interpolated to age intervals as provided by [43]. **Bottom:** Extrapolated, pandemic contact patterns 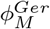 for simulated lockdown phase for Germany (as of end March in the United Kingdom; based on contact study [45]).

#### 3.2.2 SARS-CoV-2-related minimum patterns

The potential of possible contact reduction is limited by the minimum number of necessary contacts that keep essential sectors of the society running. To assess this, we consider the contact study [45], that started during the lock-down phase in the United Kingdom. By the end of March, many ‘non-essential’ parts of the economy were shut down and social interaction was limited to a minimum [40]. This study yields the minimum contact matrix 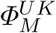.

The matrix 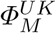 is missing values since only individuals aged 18 or older participated in [45]. In order to fill out the missing information, we follow a strategy similar to [56]. We employ the pre-pandemic/baseline contact matrix 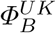 from [68]. We scale this matrix by the ratio of the dominant eigenvalues *λ*_*B*_ and *λ*_*M*_ of the lower-right, square matrices 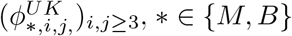. Then, we use this scaled version to fill out the missing subset

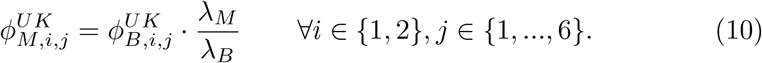

We aim at deriving a minimum contact matrix 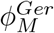 for a simulated strictest lockdown in Germany. We consider the number of contacts in the UK by the end of March to be a minimum that we can achieve in a SARS-CoV-2-related lockdown. From the UK data, we consider the quotient of contact reduction

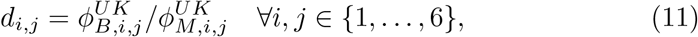

and apply these factors to the matrices 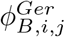 derived from [68, 37]

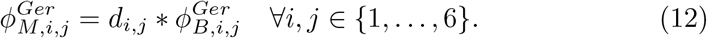

In Figure 4, the minimum 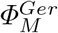 is shown at the bottom.

Note that the single entries given in the bottom row of Figure 4 are not required to be smaller than the ones in the top row. The minimum of contacts during lockdown is to be understood as the minimum of total contacts of all individuals. Locally, for one location and the interaction of two age groups, the mean contacts could even increase slightly. For example, we see a small increase of home contacts between individuals older than 80 years (factor 1.28), while there is a drastic decrease of home contacts of individuals aged 80 years and older with individuals between 15 to 35 years (factor 9.39). To conclude, the difference between the top and bottom of Figure 4 defines realistic boundaries for all non-pharmaceutical interventions that could possibly be implemented. To assess uncertainty, we allow for a 5-10 % deviance of the given values in our ensemble runs.

From [26], we have an estimated contact reduction during spring lockdown in Germany of 63 %, taking the minimum values here, we could achieve a contact reduction of 76 %.

### 3.3 Contact-related interventions

In this section, we discuss a variety of NPIs aimed at reducing the contacts between individuals.

#### 3.3.1 General interventions

There are two ways to reduce potentially dangerous contacts, namely, to avoid the contacts at all or to wear masks *and* keep distance. The latter combination has a significant effect. In many studies such as [88, 8], face coverings and ejected air flows while breathing, speaking, or coughing are studied. The meta analyses in [60] and [70] find (large) protective effects of face masks for SARS-CoV-2 transmission such as 40 % or even a pooled odds ratio of 0.35. In particular, the protective effect of community-wide masks is shown in [25]. We will consider different risk reduction ranges for wearing masks combined with keeping distance and regular ventilation of closed spaces. We also take into account that face masks are still more often worn in business than in private situations [7].

In the predictive analysis of the spread of infectious diseases, not only non-pharmaceutical interventions and one-time contact changes but also adherence to interventions is important. While there is a small decrease in adherence to preventive measures observed in [47, 7] after months of the pandemic, the adherence is still large and rather stable with an even increasing number of people wearing masks [7] (e.g., 93 % wear them often or always). In the consequence, we do not include these opposing effects in our simulations.

#### 3.3.2 Work-related interventions

While the mean number of daily contacts in Germany is lower than in many other European countries, a relatively large percentage of contacts happens at workplaces [62]. Hence, many transmissions can be avoided by working from home whenever possible.

From a recent analysis of Germany [12], up to about 50 % of the population could work from home if necessary. We suppose that in the pre-pandemic phase “home office” was only used by 5 % with as much as 25-35 % working from home during different phases of the pandemic [12, pp. 96-101]. With *Φ*_*B,W*_ the pre-pandemic contact patterns at work (Fig. 4, top) and *Φ*_*M,W*_ the minimum contact patterns in simulated lockdown, we can achieve an average contact reduction of up to *r*_*W*_ (*Φ*_*B,W*_ *− Φ*_*M,W*_) with *r*_*W*_ *≈* 0.5.

Further contact reduction is induced by people who stop working altogether, so these values have to be subtracted from the pre-pandemic matrix. We know that about 20 % of the population stopped working in March and April [12, p. 96]. For the less strict interventions, we assume values of 5-10 %.

#### 3.3.3 School-related interventions

While working from home is feasible for a larger part of the population, global school closures and the resulting home schooling “present an unprecedented risk to children’s education, protection and well-being” [86]. Apart from school closures, contacts in schools can be reduced by smaller classes where possible, fixed seating arrangements, regular ventilation, pooled testing [84, 31], and the general interventions in Section 3.3.1. We will simulate different degrees of contact reductions in schools.

#### 3.3.4 Other location-related interventions

There are a number of further locations where contact reductions are feasible such as bars and restaurants. Restrictions are also possible for essential locations like supermarkets or in public transport. To account for this, we will simulate different scenarios combined with general interventions such as masks and distancing and regular ventilation of closed spaces.

### 3.4 Deducing contact patterns from interventions

In this section, we provide the influence of the NPIs on the contact patterns and commuter rates. Let *ϕ*_*B,∗,i,j*_ and *ϕ*_*M,∗,i,j*_ denote the mean daily contacts as shown in Figure 4 in the top and bottom row, respectively, with 1 *≤ i, j ≤* 6 for the age groups and *∗ ∈ {H, S, W, O}* for the locations.

Since different interventions like working from home and face masks reduce the mean contacts by sequentially applied multiplicative factors to the remaining contacts, we use a multiplicative definition. For two intervention levels *l*, e.g., *l* = 1 for gathering bans and *l* = 2 for face masks and distancing, we introduce factors 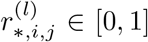 with *l* = 1, 2 and *∗ ∈ {H, S, W, O}*. We define the resulting contact patterns as

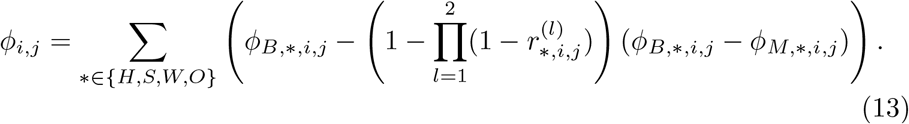

The factors 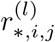 corresponding to each intervention are summarized in Table 3. If all factors are set to zero, the baseline contact rate is obtained.

In principle, for the modeling of large events such as carnival or Oktoberfest, we could also increase the contact rates by setting 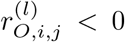 for example, taking part in carnival activities raised the infection risk from 9.5 % to 21.3 % [83]. With the current spread of the disease, these events, however, are unlikely to take place and are not considered here.

Note that a third level could be used to, e.g., implement particular awareness of senior population. In our simulations, however, including additional parameters for senior contact reduction always led to underestimations of the number of deaths.

## 4 Epidemiological parameters

In this section, we provide interval ranges for the epidemiological parameters used in our model and lay out how to implement important parameters such as testing-and-tracing limits.

The main references for each parameter are given in the corresponding subsection. For certain parameters, we also refer to the derivation in [53]. Furthermore, we derive different parameters in the clinical section of our model (stages and transitions between *I, H, U, D*, and *R*) from the LEOSS study [58]. Some recent results from LEOSS can also be found in [44]. Based on 3 265 hospitalized patients with Covid-19, we provide age-specific mean values and confidence intervals for the probabilities 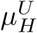 and 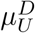. For the time spans 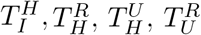, and 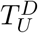, we use median values and uniform distributions.

Further, we also use provide distribution fits to the age-resolved time span data. We considered *gamma, Weibull, lognormal, normal*, and *exponential* distributions and fitted according to the lowest Pearson’s cumulative test statistic. The distributions and their parameters are given in the appendix. Note that the described distributions are of larger interest when using agent-based models.

### 4.1 Transmission risk, secondary attack rate

After the number of contacts, the most important factor for the spread of the disease is the transmission risk *ρ*_*i*_, *i ∈* 1, …, 6. As mortality is clearly found to be age-specific for Covid-19 [59], age-dependent susceptibility or transmission is less clear and under discussion [95]. There is, however, evidence showing odds ratios smaller than 0.5 for children below 15 years compared to adults [93, 28, 36], with an odds ratio of about 0.34 for children compared to adults and 1.47 of seniors above 65 years compared to adults. The meta analysis of [57] also showed higher susceptibility in adults than in children.

From [41], we find the number of second generation infections divided by the number of close contacts of the first generation as 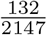. From [80], we know that the secondary attack rate in households was found to be lower than 20 % (in most of the studies) and thus even lower in general (non-household) settings. From [11] and the first cases of Germany, we have a secondary attack rate of 5-10 %. The meta analysis in [57] found a pooled secondary attack rate in household settings of 18 % and of 0-5 % in workplace, school or social settings. Taking relative susceptibility as described before into account and adding uncertainty, we have 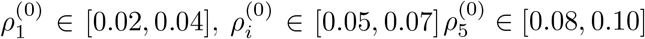. From the mortality for Germany, we derived that a slightly larger 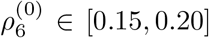 had to be used to account for many infection clusters in residential homes for the elderly [73]. Then, we define

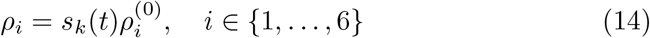

with the seasonality factor *s*_*k*_(*t*) given in the next section in eq. (15). By this parameter, we take the influence of seasons into account, a factor that is still under discussion in the literature [81, 24]. We will allow for a slight variation of the infection rate with the seasons as detailed in the next subsection.

### 4.2 Influence of the seasons

As Covid-19 raged all over the world and did not disappear in the summer months, only reduced seasonality can be expected. While [24] found that multiple meteorological aspects such as temperature, humidity, and wind speed influence the spread of the disease, the analysis of over 100 articles in [81] found only a “weak modulation effect” so far, with more evidence only to be expected in 2021.

However, another important aspect of seasons is the transition from outdoor to indoor contacts. While counter-strategies like face masks [88, 70, 25] or adapted air flows [8], or both [21, 20] are studied, it is reasonable to assume that they cannot fully waive the risk of indoor infections. In [69], the large majority of considered clusters appeared in closed environments, while [65] even quantified the risk of indoor infections 18.7 times larger than the risk of open-air infections. Additionally, [21, 20] gave particular “maximum exposure times” for different indoor settings. As we do not distinguish between contacts in closed spaces and in outdoor spaces, we allow the infection rate *ρ* (cf. (14)) to vary slightly with the seasons by using

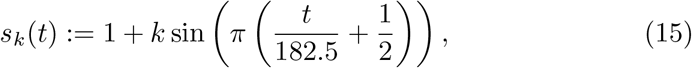

where *t* is the day of the year and *k ∈* [0.1, 0.3]. The chosen parameter *k* will yield scenarios with modest seasonal influences. From the simulation results, we will see that the integrated seasonality is capable of reproducing low and only modestly increasing numbers of infections to the end of the summer period given only mild restrictions on contact reductions. On the other hand, we can also correctly model the recent horizontal development of detected cases with stricter NPIs in autumn and beginning of winter scenarios. For more details, see Section 5.

### 4.3 Quarantine and Isolation

Note that we use the parameter 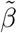(or 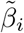) differently than the *β* in the most recent version of [53]. In the latter, *β* represents the risk of infection from the detected and infected symptomatic patients not yet effectively isolated. It varies within countries, cultures and healthcare systems and is assumed to be in the range of [0.05, 1.0] [53].

The parameter 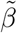 in our model additionally represents the share of infected people *I*_*X*_ that are not detected. For comparison, suppose *I* = *I*_*X*_ + *I*_*H*_ + *I*_*R*_, i.e. the infected are the sum of the undetected *I*_*X*_, those later hospitalized *I*_*H*_ and those that recover on their own *I*_*R*_. Then, our 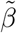 and *β* in [53] are related by

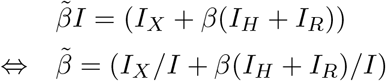

A value of 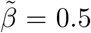 could then mean that the number of unreported cases *I*_*X*_ equals the number of reported cases *I*_*X*_ = *I*_*H*_ + *I*_*R*_ = *I/*2, and reported cases are *perfectly* quarantined from the first moment of infection (*β* = 0) while nonreported cases move freely for the whole time of infection. Of course this is only the very unlikely edge case. In reality, detected individuals are neither detected nor isolated from the very point of symptoms onset while also undetected individuals may isolate themselves due to their symptoms.

The decrease of 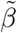 is what the national strategy [71] of test, trace, and isolate majorly contributes to [27]. Unfortunately, we already faced “no longer traceable transmission chains” [74]. Since it is difficult to quantify how phases of relative transmission control or “diffuse spread” [74] influence 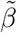, we assume 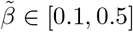 and consider different scenarios. Thereby, we let 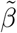 increase along a sigmoidal (cosine) curve from the minimum in [0.1, 0.3] to the maximum value in [0.3, 0.5]. We assume the minimum value for phases with less than 50 infections per 100 000 individuals increasing smoothly to the maximum value for cases of about 250 infections per 100 000 individuals.

#### 4.3.1 Exposure, carrier, and infection transition

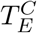 denotes the time elapsed between exposure and carrier state and 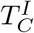 the time between the beginning of carrier state and the onset of symptoms. We assume that these parameters do not depend on age. 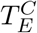 represents the latent period, namely the time an individual remains in a latent non-infectious stage following the transmission. The incubation period is the time between exposure and when symptoms are first apparent, i.e.

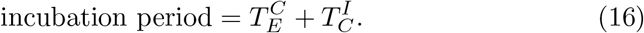

We use an incubation period of 5.2 days and assume that infections occur randomly during the infectious period [53]; for more details, see [53, 64, 94]. We allow for 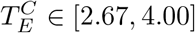. Hence, by median, 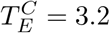 and 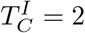.

### 4.4 Asymptomatic individuals

During the early outbreak in Germany, a proportion of 22 % of infections was reported to be asymptomatic [83]. Two recent meta analyses [22, 14] found ranges of asymptomatic cases from 3-67 % with an overall estimate of 17 % and 20 %, respectively. The role of children is still heavily under discussion, and the number of asymptomatic cases in children is even less clear [48, 22, 28, 95]. For instance the studies [28, 95] considered children diagnosed with SARS-CoV-2 infection. The former found 28 % of asymptomatic cases compared to 12 % in adults, and the latter found 19 % with another 25 % of mild cases. We thus consider 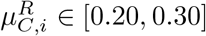 for *i* = 1, 2 and 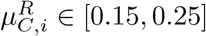 for *i >* 2.

While the asymptomatic individual remains infectious, we assume

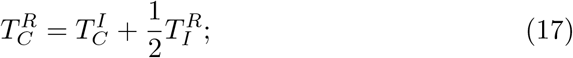

for more details, we refer to [53].

### 4.5 Symptomatic individuals

Symptomatic cases are divided into those with mild symptoms not requiring hospitalization and those where an initially mild stage becomes severe. From [72], we have a range of 5 % to 22 % of hospitalized cases with respect to the reported number of cases in Germany. The highest number in the report occurs during a time of presumed huge underreporting and when also many elderly people were infected. In [87] and its adjusted version for Great Britain [34] the symptomatic cases range from about 0 % for age group 0-9 to 18 % and 27 %, respectively, for people older than 80 years. We range our samples accordingly and allow for a deviance of up to 20 % from the mean or median; see Table 2.

For the recovery time 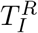 of mild cases, we take a median of about seven days [90, 53] With a deviance of up to 20 %, we consider a range 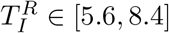. For the time span 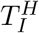 of initially mild symptoms becoming severe and where hospitalization is needed, we assume 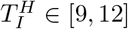 for patients aged 0-34 years and 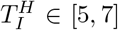 for individuals older than 35 years. In [53], we refer to some earlier studies [32, 54, 42] providing a median period of 4, 4.9 and and 7 days, respectively. From [58] and 3 265 patients, we have a median *time of known infection to hospitalization* of 9 days for age group 0-14 and 12 days for age group 15-34, and 6 days for 35-65 years, 7 days for 65-75 years, and 5 days for 75+ years; see Figure 16. Note that the time of known infection, however, only provides a lower bound for the time of symptoms onset to hospitalization.

### 4.6 Hospitalized individuals and and transfer to intensive care

In our model, there are essentially two possibilities for patients which are admitted to a hospital. Either individuals recover directly in the hospital or individuals are transferred to intensive care units with probability 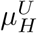.

In an early study [89], 26 % of hospitalized patients needed intensive care which comes close to the 25 % reported in [50]. In our analysis of LEOSS [58], we see an increase from no cases or only a few for the youngest individuals to 37 % for the age group 66-75 years, with a surprising slight decrease to 24 % for the age group 80+. In contrast, [34] provides a monotonous curve from 5 % to 70 %. Combining the results, we let 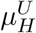 range from 0.05 to 0.45 for the different age groups; see Table 2 for details.

The median time from symptoms onset to intensive care 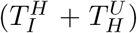 is estimated to be 9 and 9.8 days in [32, 54] (with a median of 4 and 4.9 days from symptoms onset to hospitalization, i.e., 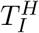). From [58], we derive hospital stays for patients admitted to ICU (and recovering later on) of 11.3 to 8.4 days on average (median 13 to 5 days). This includes, however, days before and after intensive care; see Figure 17. Combining these results, we consider a range of 3 to 7 days for 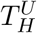 for all age groups.

For the time 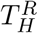 of individuals recovering in the hospital without needing intensive care, we refer to our reasoning in [53] where 7-16 days were considered. From [58] and Figure 18, we have median recovery time spans of 4 to 6 days for age groups 0-14 and 15-35 years, respectively. This value rises to 8 days for 36-65 years, 10 days for 66-75 years, and 15 days for 76 years and older.

### 4.7 Intensive care, recovery and death

At last, we have to determine the mortality 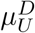 of patients in intensive care and the patients’ average time spans to either recovery 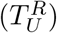 or death 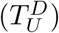.

For 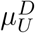, we use the ranges as presented in Table 2 based on a combination of the following findings: In [4], the overall ICU mortality is estimated as 41 %. The mortality among mechanically ventilated patients is considered in [50] and ranges from 28 % to 72 % from the youngest to the oldest age group. For non-ventilated patients, these numbers range from 1 % to 34 %. From the LEOSS survey [58], we have ICU mortalities of 0 %, no cases, or large confidence intervals around low values for the youngest individuals, increasing up to 57 % for patients aged 76 years and older; cf. Figure 6.

**Fig. 6.**
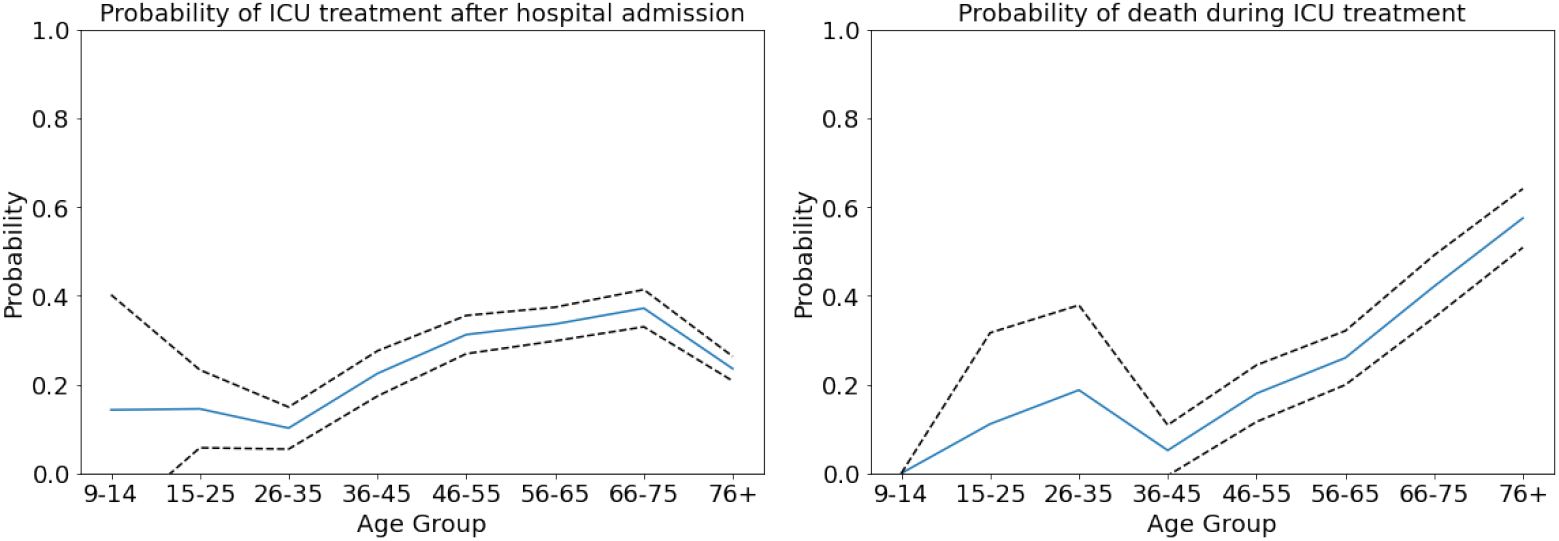
Graphs derived from LEOSS [58]. Mean value shown in blue, 95 % confidence interval shown in dashed black lines. **Left:** Age-specific probability 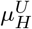 to need intensive care once hospitalized. **Right:** Age-specific probability 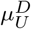 to die once intensive care was necessary.

The recovery time 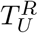 for severe or critical cases can take up to 3-6 weeks [92, 53]. In [54], the pooled median from symptoms onset to recovery was 18.3 days. However, this also potentially includes mild cases. From the LEOSS survey [58], we have mean recovery time spans of 6 and 11 days (median: 7 and 7 days) for people between 0-14 and 15-35 years. For older people, the average time spans are 17, 21, and 15 days (median: 14, 18, and 9 days); see Table 2 for the ranges used in our simulations.

The pooled time span 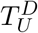 from symptoms onset to death is estimated as 15.9 days in [54]. In our database from [58], no death is observed for individuals aged 0-14 years. For the age groups, 15-35, 36-65, 66-75, and 76 and more years we have mean values of 12, 18, 18, and 12 (median: 15, 16, 15, 9). In [53], we also assume 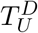 to be slightly shorter than 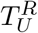. For the ranges finally considered in our simulations, see Table 2.

## 5 Results

In this section, we present different scenarios based on the course of the pandemic in Germany. Four of these scenarios are retrospective and start at June 1, July 15, September 1, and October 10, respectively, and they last for a period of 45 days. Twelve scenarios are considered for the time span of December 12 to January 10, in particular focusing the Christmas holidays. Our aim is to qualitatively capture the trend of infections and their time-dependent distribution across Germany. We use the same set of epidemiological parameters for every scenario without adapting them for the specific situation. We only implement different sets of interventions that lead to different rates of contact reductions. The mean effectiveness of the sets of NPIs is derived from the development of SARS-CoV-2 infections and Covid-19-related deaths in Germany.

### 5.1 Aims and Expectations

*Our first aim is to qualitatively capture the infection trends and death rates for all scenarios* without tweaking specific parameters for a better local fit. We thus do not expect to exactly predict the number of deaths or infections for all scenarios. We rather expect slight under- and overshoots depending on the scenario and the quality of the underlying data. The simulation times in the retrospective scenarios are rather long and we know that predictions for many weeks become increasingly less reliable. We do not expect that this is different for our model. What we want to see here is that our model reproduces the main trend.

The prediction of SARS-CoV-2 infections or related deaths is a challenging task and heavily depends on the input data. Our model is age-resolved, but some of the input data is not. While already relatively good data is available from RKI [43] and DIVI [66], additional data may be necessary to further validate our simulations. For the deaths reported with age resolution, only the dates of the assumed infection are available in the repository [43], and we thus have to extrapolate the time of death from the time of infection in the real data. This is then only an approximation to the real life situation and at times this extrapolated number differs considerably from the overall number given in the daily situation reports. The overall number in the situation reports, however, is not age-resolved and not easily accessible by automation. ICU occupancy also lacks age resolution, so we have to extrapolate intensive care for the different age groups. On top of that, the underlying data that we compare to is uncertain as well. The next step would be to further vary that as well over the different simulation runs.

A further approximation in our model is that people can only die after they were in intensive care first; cf. Figure 1. In real life, and in particular in times of high infection rates, Covid-19-related deaths can also happen in, e.g., residential homes for elderly care. This leads to an overestimation of ICU occupancy in our model.

*Our second aim is to capture the regional spread of the infection across Germany*. Our model must be able to predict the spread of the virus along the routes provided by our mobility data. We want to observe that local hot spots affect their neighbouring regions over time.

Our model is only calibrated with real data once at the beginning. Hence, local effects are very hard to capture in the simulation. For example, in the real world, the implemented local NPIs in regions with very high incidence might differ considerably in their effectiveness, so that a once local hot spot will no longer be hot after some time or become even hotter. A further local driving mechanism of the infection are super-spreading events. As we do not yet include those as stochastic events in our simulation and since the location of these events is by nature random, we expect regional deviations between the simulated scenarios and the extrapolated real data that may, of course, also influence the global infection spread. However, if local hot spots are already contained in the initial data, we expect to see their spreading to the surrounding regions.

*Our third and main aim is to assess the mean effectiveness of mitigation strategies*. We implement local dynamic NPIs to control hot spots with more than 200 infections per 100 000 inhabitants. These are used carefully to avoid overfitting and to not distort the image of the country-wide mitigation strategy. By variation of the country-wide contact reduction over a large number of simulations runs, the comparison with the extrapolated real data will indicate how the implemented mitigation strategy influences the effective contact reduction.

### 5.2 Retrospective Scenarios

For all simulations, we use the ranges for the epidemiological parameters in Table 2. We run sets of 1000 Monte Carlo runs with different levels of mitigation as available from Table 3. We then identify ranges for the effective mitigation in the different scenarios by comparing our simulation results to the real data.

In all scenarios, school holidays are implemented for the corresponding dates for all federal states. Other than that, we group the most relevant interventions on state level to single dates and consider their mean effectiveness over periods of multiple weeks in order to avoid overfitting. In the following, for better readability, we drop the indices *i* and *j* of the NPI-related parameters 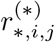 cf. Table 3.

#### Set of NPIs for June to October

For the time span of June 1 to September 30, we model rather weak effective contact reductions – in accordance with the lifting of many interventions in the summer period as described in [17, 15] – to reproduce the slow spread of the disease. We assume contact reductions in the range of 0 to 20 % in home and other locations, a working from home percentage between 20 and 30 %, up to 5 % of people who stopped working altogether (also cf. [12]) as well as an additional contact reduction in all locations by wearing masks and distancing of up to 20 %. These considerations translate to 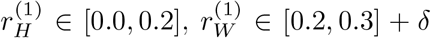 with *δ ∈* [0.0, 0.05], 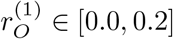 with 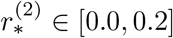, *∗ ∈ {H, S, W, O}*; see Section 3.3.1.

In addition, by averaging we implement the partial school closures and remote schooling 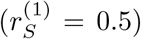 from June 1 until June 15 for all states; after that, schools operate as usual with 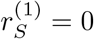 (except for the school holidays on federal level, where 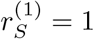.

From our simulations runs, we assess the effectiveness of the mitigation strategies in this period on the overall contact reduction by 15 % with schools open and by 30 % during school holidays.

#### Set of NPIs for October

From October 1 to October 30 we assume slightly stricter contact reductions than in the summer based, e.g., on the call to further attentiveness end of September [19]. We identify slightly increased values for contact reduction at home (20 to 40%, i.e., 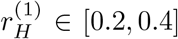) with increased values for protective interventions such as wearing masks, regular ventilation of closed spaces, or distancing (20 to 40%, i.e., 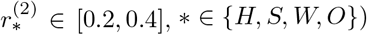. All other NPIs are kept in place as before. The NPIs’ effect on the median overall contact reduction that we obtained from our simulations is about 31 %.

#### Set of NPIs for November

For the time span of November 1 to November 30, stronger restrictions are in place [18]. We assume contact reduction in homes by 40 to 60 %, a proportion of non-working individuals of 0 to 10 % plus a proportion of home-working individuals of 20 to 30 %. Since restaurants, bars, and most leisure-related facilities were closed, we model a contact reduction in other locations by 60 to 80 %. Additional protective effects to avoid contacts are estimated as 20 to 40 % in homes and schools (since face masks are less worn in private situations [7, 45] and since school contacts are more difficult to reduce than work or other contacts) and 40 to 60 % in work and other situations. Through our simulations runs, we assess the effectiveness of the mitigation strategies in this scenario on the overall contact reduction with 50 %. This is slightly larger than the estimation of 43 % by [26] but still far from the reported reduction in spring (63 %) that may have been necessary to substantially mitigate the spread of the virus.

To summarize, we set 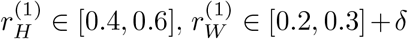 with *δ ∈* [0.0, 0.1], and 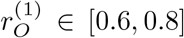. We further assume 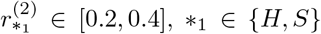 and 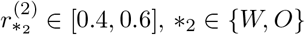.

#### Dynamic and local non-pharmaceutical interventions

In order to mitigate the spread in local hot spots, we dynamically implement a set of strict non-pharmaceutical interventions. Once 200 infections per 100 000 inhabitants and county are reached, an enforced contact reduction is implemented for 14 days. The parameters of this local set of interventions are the sames as the *NPIs for November* described above with the following modifications: 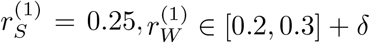 with *δ ∈* [0.1, 0.2], since additional facilities may be closed on a local scale, and 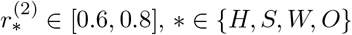, which is a quite strong measure for distancing, face masks and other interventions.

#### Simulation

Our initial conditions are derived from the age-resolved case data provided by [43]. We take confirmed cases around the start date of our simulation from which we extrapolate the compartments in eq. (2)-(9) by using the parameters in Table 2. Based on [66] and our parameters, we extrapolate age-resolved ICU data. In order to obtain age-resolved ICU data, we slightly reduce the initial extrapolation of 80+ intensive care cases since too large death rates occur in the beginning part of the simulation otherwise. We refrained from further correction of initial extrapolations without having more reliable data.

Given positive rates of 1 % or less during the summer [75], we assume that the number of unknown symptomatic infections was very small. Therefore, we start our simulations on June 1, July 15, and September 1 directly from the number of confirmed cases. Given the increased proportion of positive tests up to mid of October, we start the the simulations with a twofold of the confirmed cases. For each scenario, we run 1000 Monte Carlo runs such that we have a reliable set of parameters sampled in the given ranges. For the runs, we provide the median values as well as the percentiles obtained from the simulation runs. We use a seven day moving average of real world data and extrapolate the day of death, using the parameters from Table 2.

### 5.3 Discussion of Retrospective Scenarios

In the following, we discuss the results of the four different retrospective scenarios with the described sets of implemented NPIs over time. We compare the overall and the age-resolved death rates with the extrapolated real data. We also compare the overall infection rates and the ICU occupancy. In the corresponding Figures 7, 9, 11, and 13, we present the median (percentile p50) and the percentiles ranges from (p05 and p95) as well as p25 and p75 as explained above. Additionally, in the maps of Figures 8, 10, 12, and 14, we show two snapshots of the regional spread of the infection, where we compare the extrapolated real data on the left with our simulation on the right. Note that the scaling of the color bars differs for the scenarios and represents the relative number of infections per 100 000 inhabitants.

**Fig. 7.**
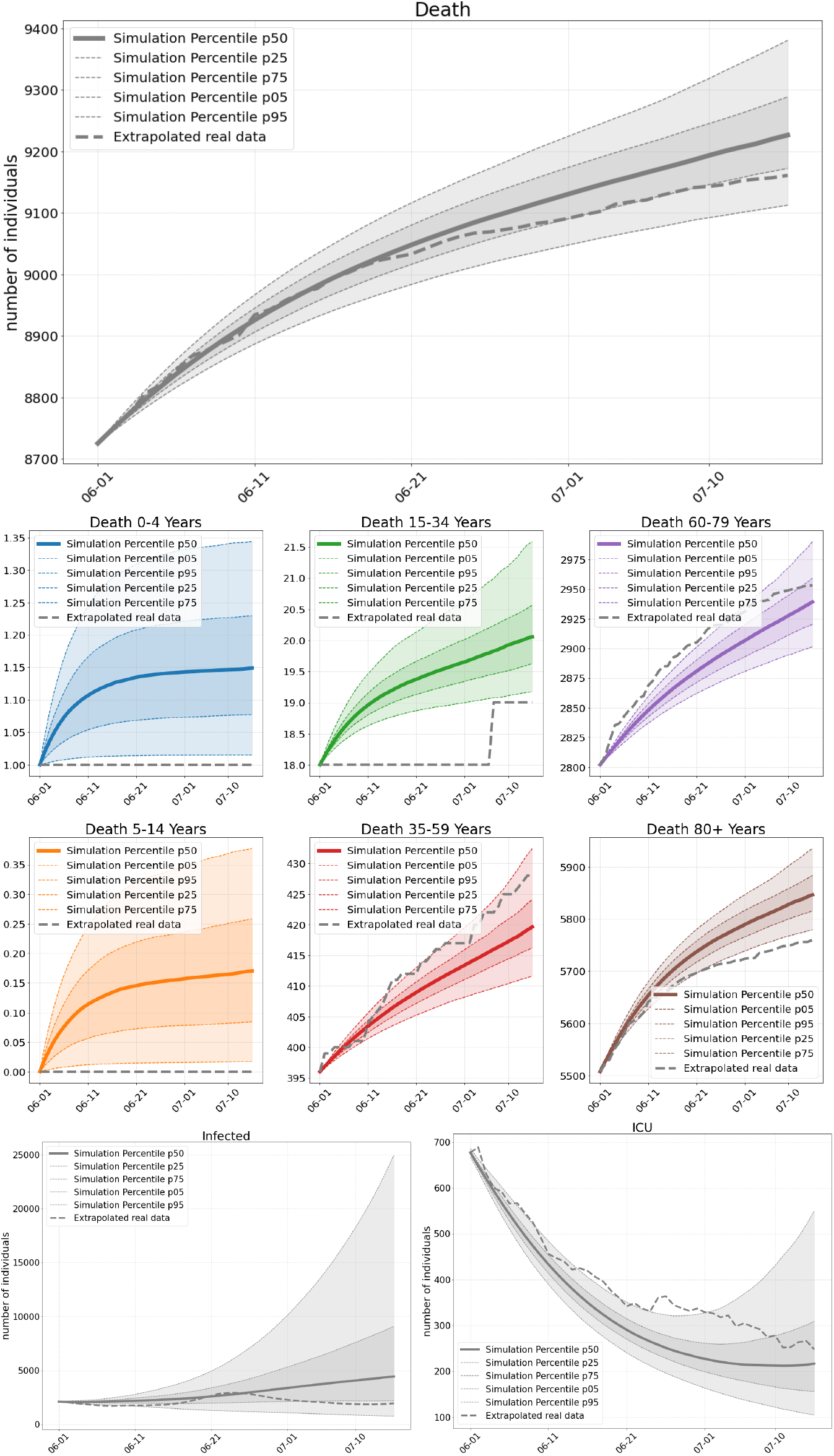
S1: Total number of deaths (top), and deaths per age group (center), total number of SARS-Cov-2 infections (bottom left) and occupied ICU beds (bottom right) as of our simulations compared to extrapolated real world data from July 15 to August 29. On average we have 0.4 exceedances of 200 infections per 100 000 inhabitants where stricter local NPIs are implemented.

**Fig. 8.**
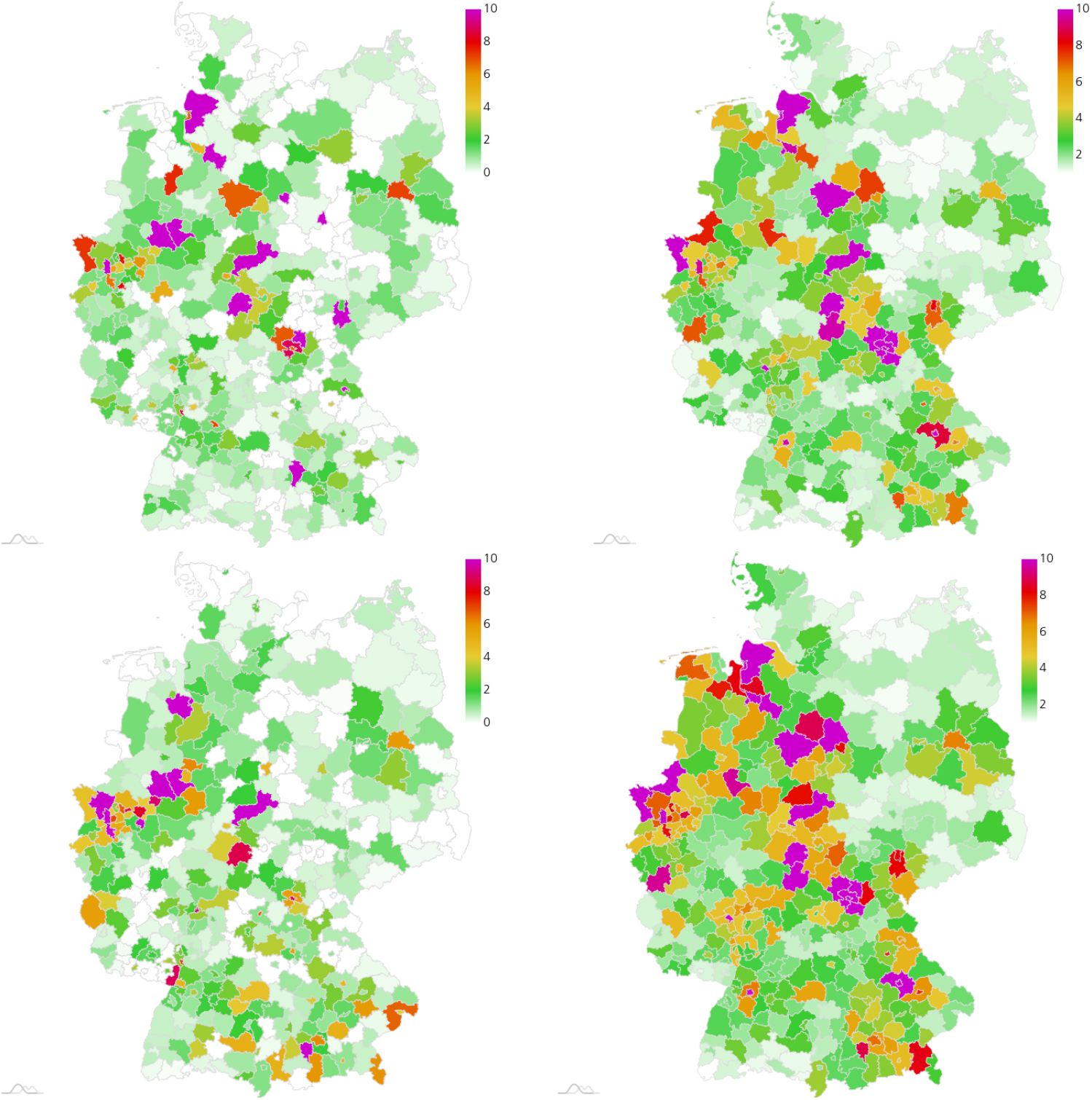
S1: Extrapolated real world data (left) and simulated number of infections (right), relative per 100 000 inhabitants, on June 15 (center), and June 30 (bottom). Maps created with amCharts.

**Fig. 9.**
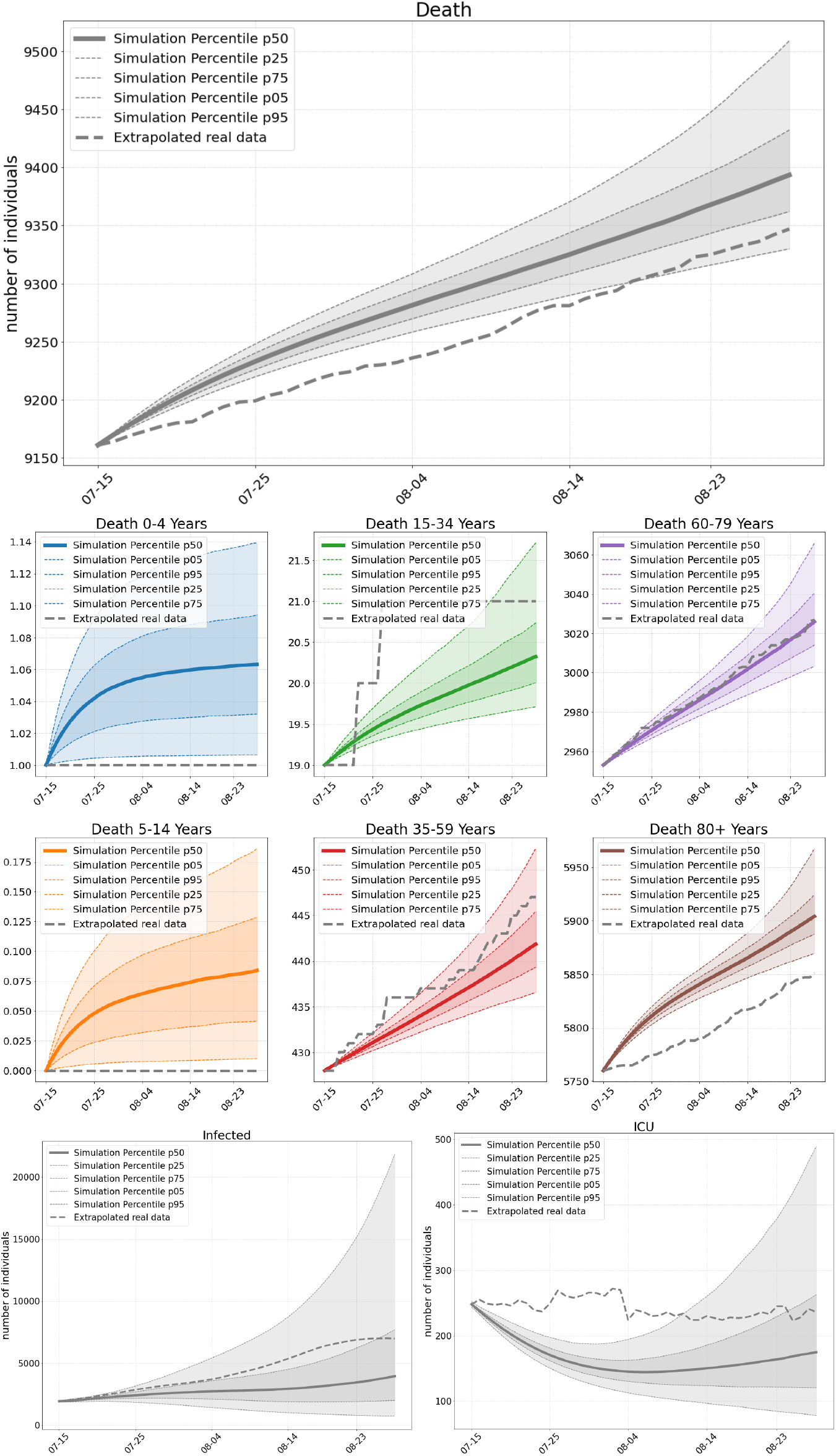
S2: Total number of deaths (top), and deaths per age group (center), total number of SARS-Cov-2 infections (bottom left) and occupied ICU beds (bottom right) as of our simulations compared to real world data from July 15 to August 29. On average we have 0.03 exceedances of 200 infections per 100 000 inhabitants where stricter local NPIs are implemented.

**Fig. 10.**
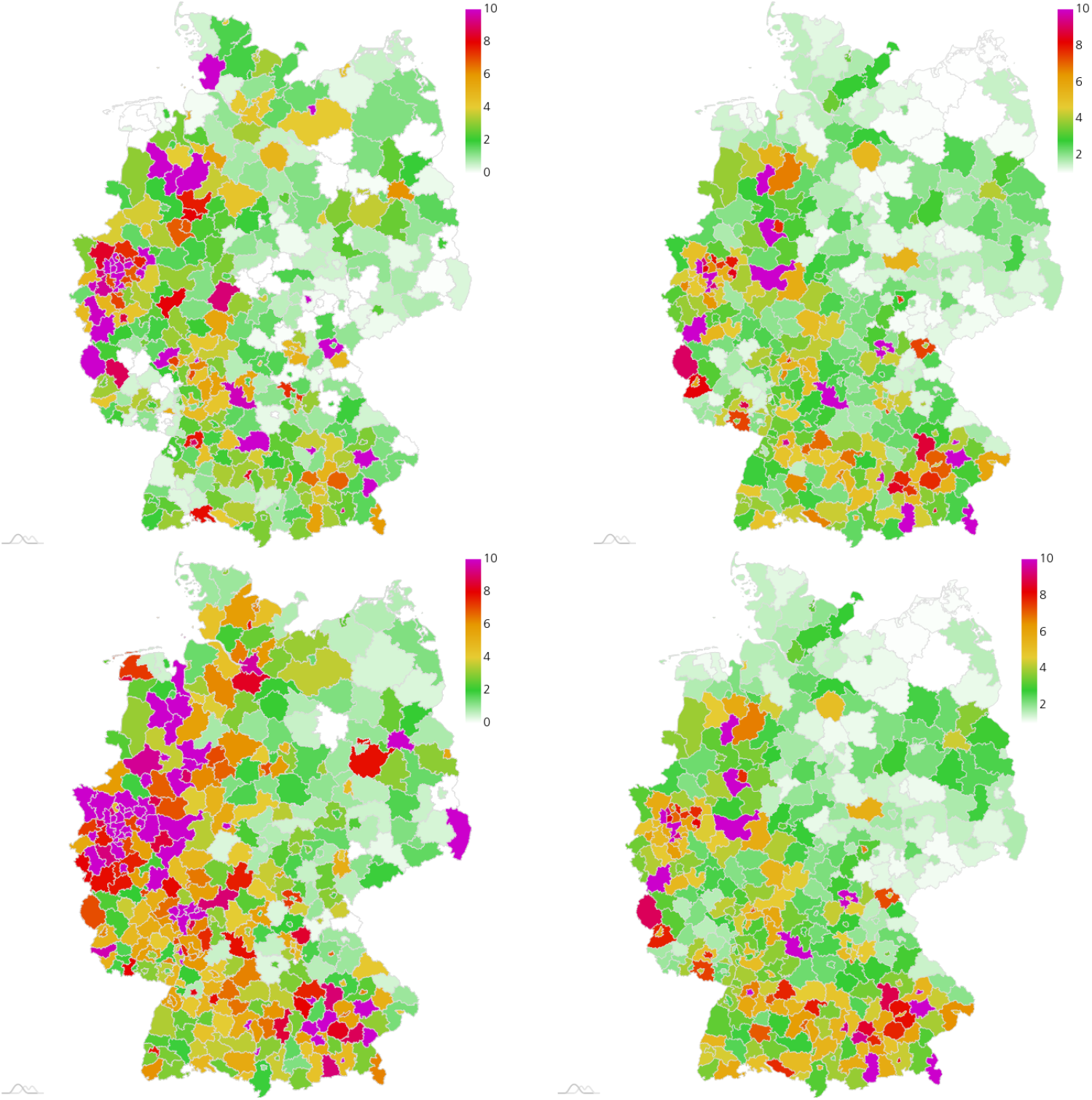
S2: Extrapolated real world data (left) and simulated number of infections (right), relative per 100 000 inhabitants, on July 30 (center), and August 14 (bottom). Maps created with amCharts.

**Fig. 11.**
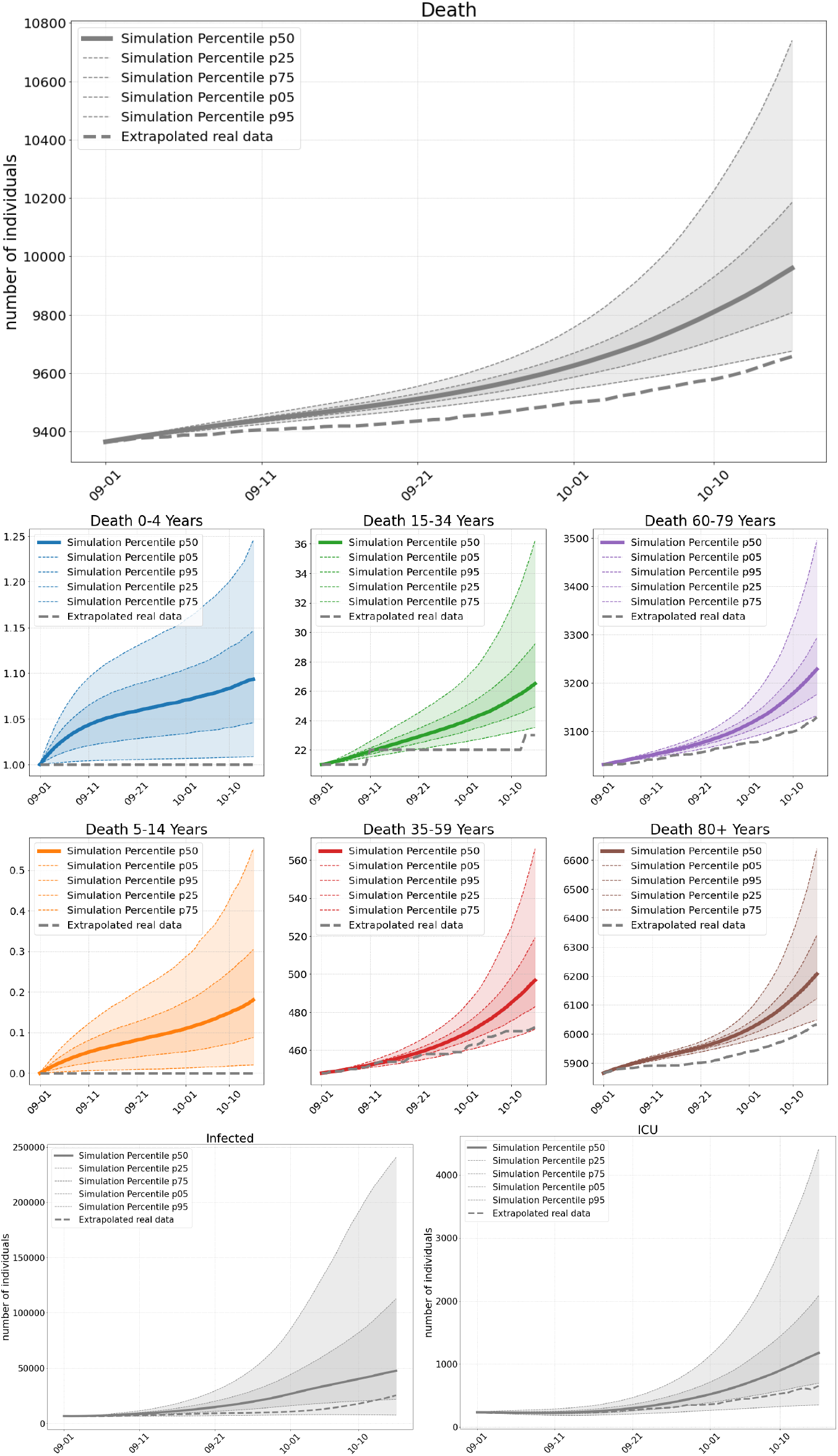
S3: Total number of deaths (top), and deaths per age group (center), total number of SARS-Cov-2 infections (bottom left) and occupied ICU beds (bottom right) as of our simulations compared to extrapolated real world data from September 1 to October 15. On average we have 44 exceedances of 200 infections per 100 000 inhabitants where stricter local NPIs are implemented.

**Fig. 12.**
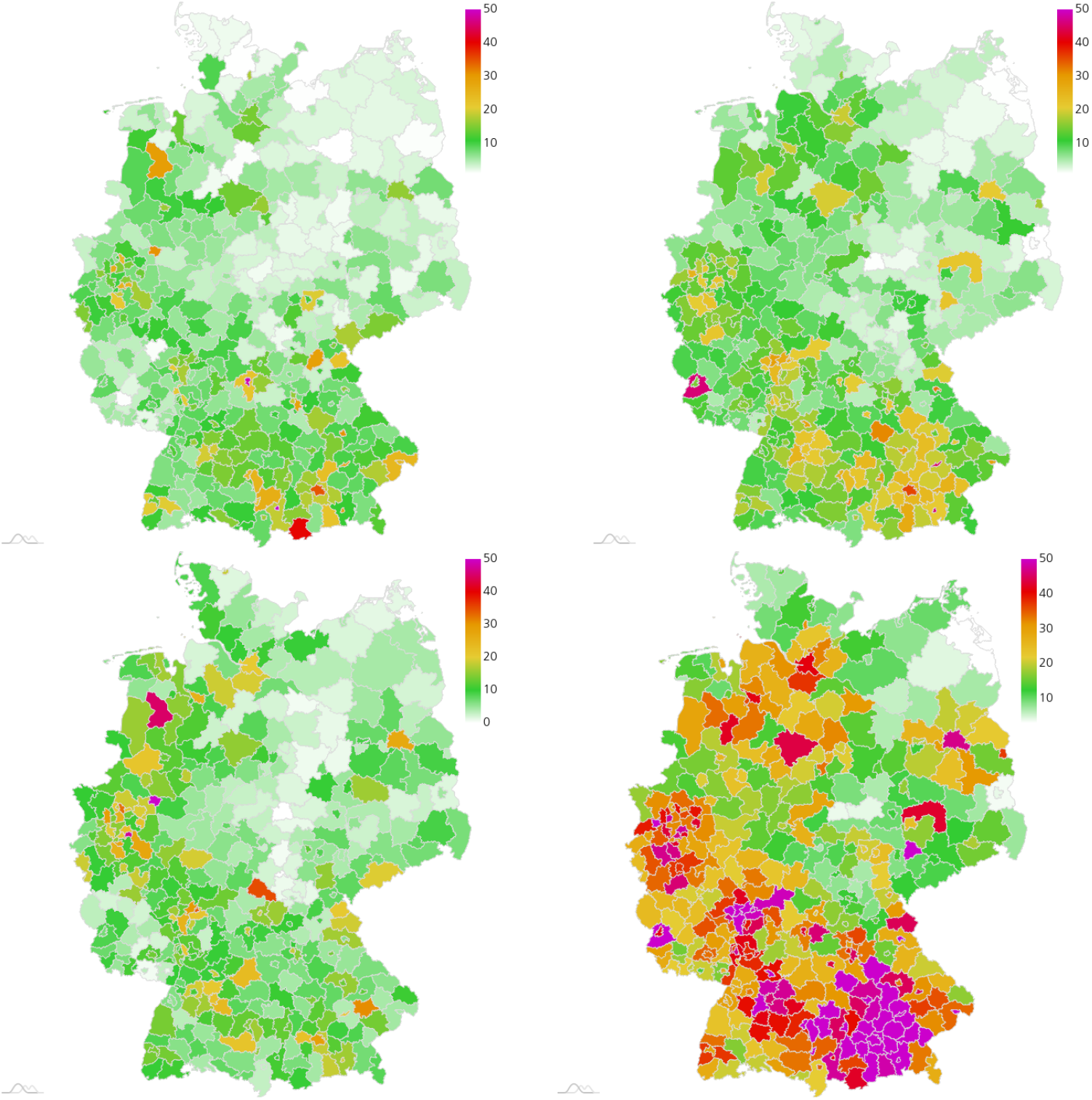
S3: Extrapolated real world data (left) and simulated number of infections (right), relative per 100 000 inhabitants, on September 15 (center) and September 30 (bottom). Maps created with amCharts.

**Fig. 13.**
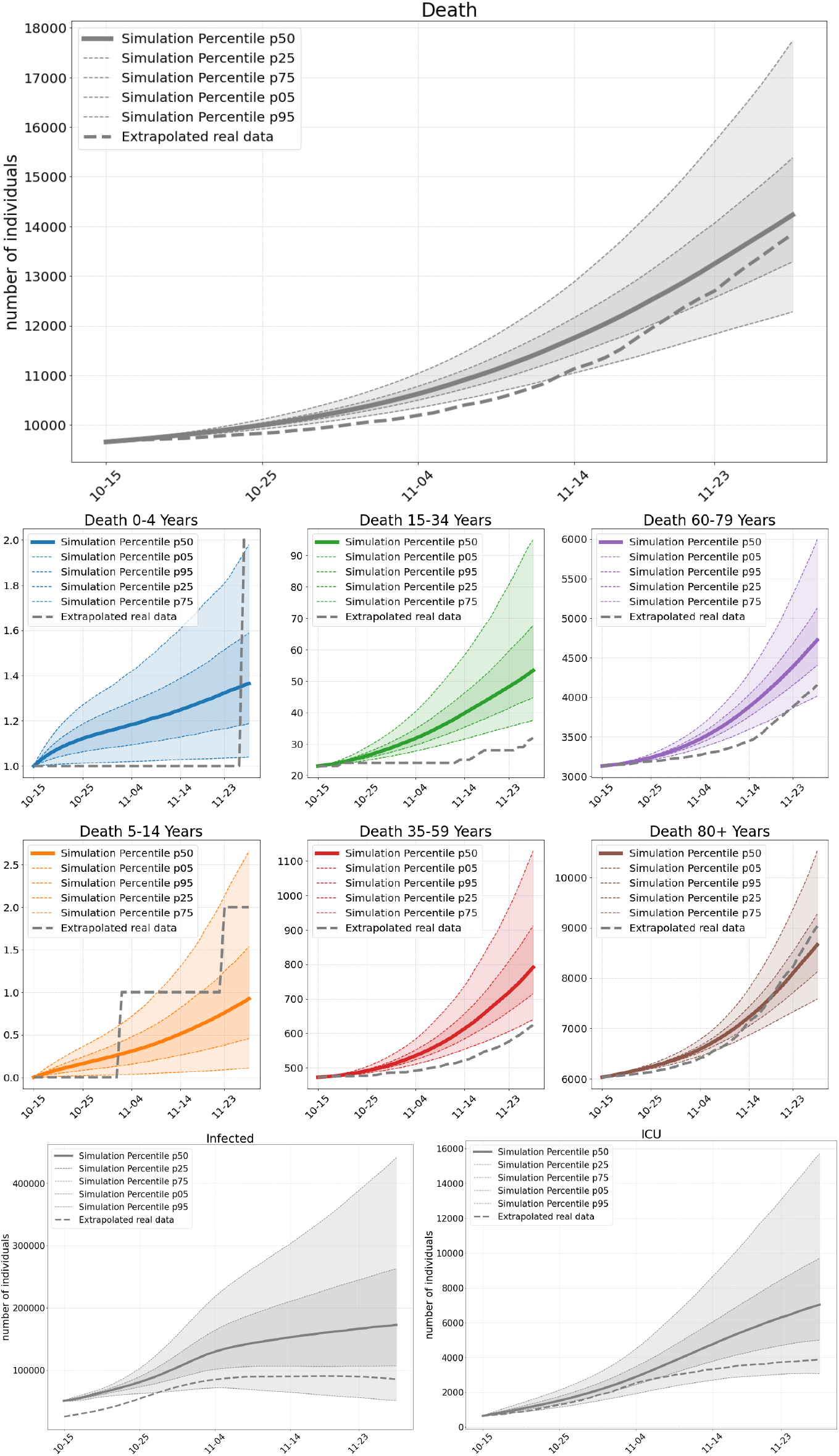
S4: Total number of deaths (top), and deaths per age group (center), total number of SARS-Cov-2 infections (bottom left) and occupied ICU beds (bottom right) as of our simulations compared to extrapolated real world data from October 15 until November 30. On average we have 371 exceedances of 200 infections per 100 000 inhabitants where stricter local NPIs are implemented.

**Fig. 14.**
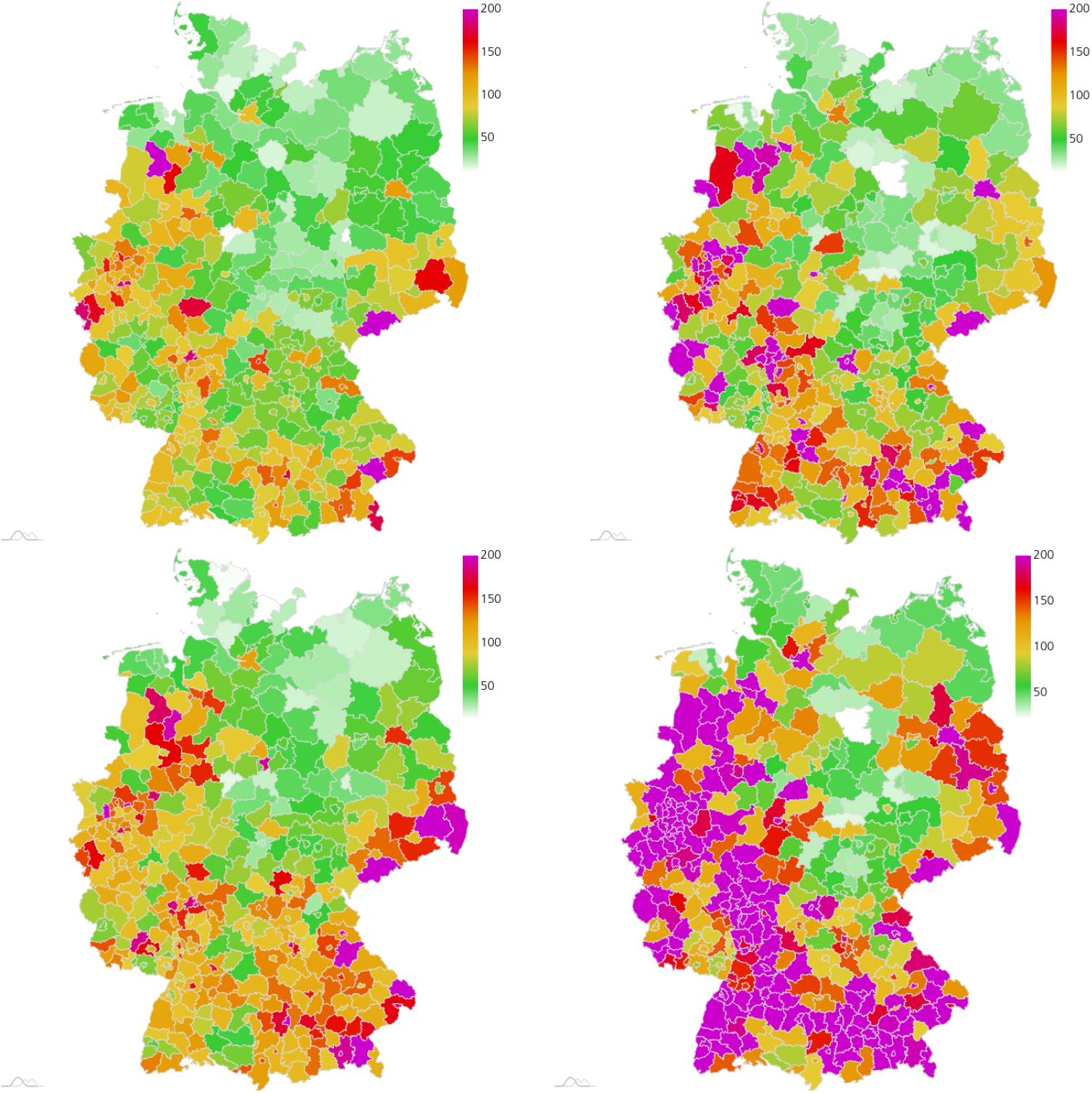
S4: Extrapolated real world data (left) and simulated number of infections (right), relative per 100 000 inhabitants, on October 30 (center) and November 14 (bottom). Maps created with amCharts.

#### 5.3.1 Scenario 1

Our first scenario (S1) is computed 45 days from June 1 onward. During the summer months, we observe only a slow rise in the infections in the RKI data and a decrease of ICU occupancy from DIVI (Fig. 7). Both are captured well with the median of our simulations. The small elevation in the number of infections in June is due to an outbreak of Covid-19 at the Tö nnies slaughterhouse in Rheda-Wiedenbrück. By June 20, the number of infections had exceeded 1 000. As expected, we cannot capture such a stochastic event with our model. This hot spot and its influence on the neighbouring regions is also missing in the simulation in the maps of Figure. 8.

Focusing on the first part of June (see the upper part of Figure 8), we observe further regions that are underestimated by our simulation and some where it is the other way around. Overall, however, if we omit the parts around Tö nnies, a larger incidence in the RKI data almost always corresponds to a larger incidence in the simulation data. The parts that are overestimated in the simulation can often be contributed to our mobility data. This is only an approximation of the real life mobility and the infection can only spread along those routes. Worth mentioning is the spread of infections into neighbouring regions which we can see for Berlin from the middle to the bottom. Due to the inclusion of mobility, we see that the infection spreads into the near regions in our simulation (right) as in the extrapolated real data (left). Due to the longer simulation time, the results on the bottom show further deviation from the data. The Tönnnies’ affected regions remain underrepresented in the simulation map. But again, we see an overall good agreement with the infection spread observed in the real data.

In Figure 7, we plot the median death rate, the percentiles and the extrapolated real data for the different age groups. Our model is quite close to the overall death rate. The age group 60-79 years is captured very well and the age groups below are also captured well. We are a little less close for the age group 80+ years. The deviations are to be expected since we aim for an overall good fit of our model without tweaking it for the individual scenarios. Additionally, we lack age-resolved ICU occupancy data and have to extrapolate the intensive care input data for the different age groups. This then also affects the simulated death rates. Note that the age groups below 15 years only contribute marginally to the death rate. The deviance with simulation seems large, but due to the small numbers (0 or 1), we are actually close to the real world value.

#### 5.3.2 Scenario 2

Our second scenario (S2) is computed 45 days from July 15 onward. The results of this Scenario are very similar to the results of S1. This holds for the overall death rates and the death rates in the different age groups as depicted in Figure 9. A main difference is the influence of travel returners [76] that might be responsible for the rise of infected in the beginning of August with a plateau reached at the end of that month; cf. Fig. 9. Such an influence is not considered in our simulation which contributes to the underestimation of infections. Similarly, we underestimate the number of ICU occupancy.

In addition to travellers, many smaller outbreaks in a number of administrative districts in various settings (social, industrial, etc.) are reported [76]. There is an increasing trend in 7-day incidences in North Rhine-Westfalia (NRW) and Hamburg and higher-than-average 7-day incidences in Hesse and Berlin, as well as a Covid-19 related outbreak in the Bavarian district of Dingolfing-Landau (DL) with *>* 400 cases. This is what we observe in the map of the extrapolated real world data on the left hand side of Figure 10.

In our simulation on the right hand side, NRW still seems to be a little underrepresented, but interestingly, we catch the outbreak in DL with our input data from July 15, as well as its spread to neighbouring regions that we also observe in the real data. Hesse and Berlin are regions of higher infection rates in our simulation as well. Some local hot spots, maybe due to weddings in that time, are again missed due to their stochastic nature. All in all, the qualitative regional infection spread is again captured well.

#### 5.3.3 Scenario 3

Our third scenario (S3) is computed 45 days from September 1 onward. Compared to S1 and S2, we overestimate the number of deaths for all age-groups (Fig. 11) compared to the extrapolated real data. However, by the end of this period, on October 15, we also have 9710 deaths reported in the RKI daily situation report [77]. This means that some of the infected died earlier then we could extrapolate from the assumed day of infection in [43]. 9710 deaths is then already much closer to the prediction of our simulation.

The slight overestimation of deaths is consistent with the projected infection numbers and ICU occupancy that are overestimated as well. As the proportion of positive tests grew considerably from 0.7 % to 5.6 % in the considered period, we assume that there might have been a larger number of undetected cases at the end of this scenario which are not reflected in the real data yet. As explained in Section 5.1, we expect our model to rather overestimate ICU occupancy in phases of high infection rates. This might also already contribute to the overestimation we observe in Figure 11.

Our simulations from September 1 predict larger incidences in multiple federal states like Bavaria, North Rhine-Westfalia, Berlin, Lower Saxony, Baden-Württemberg, or Rhineland Palatinate with lower incidence in the center and North Eastern part of Germany; see Figure 12 (right). Although we have a qualitatively similar picture in the extrapolated real data, the number of reported cases are substantially below the predicted values; see Figure 12 (left). On the other hand, we see that these and even much higher incidences are to be found in the real data in October in these precise regions; see Figure 14 (please pay attention to the different scales in these two figures). This further supports our assumptions that there might be a larger number of undetected cases in the infections that are not captured in the real data in Figure 12 (left).

#### 5.3.4 Scenario 4

Our fourth scenario (S4) is computed 45 days from October 15 onward. Due to the increased proportion of positive tests, we start this simulation with the assumption of twice as many confirmed cases, which is also the reason for the difference between the infected in the real data and the simulation data in the initial phase depicted in Figure 13. We see that the curve of predicted deaths is steeper in the beginning while the extrapolated real data curve becomes steeper towards the end of the scenario; see 14. As in the last scenario, the situation reports already yields 16 248 deaths at the end of November [78], but again the corresponding age-resolved data that we need to compare to is not directly available. In this scenario, we overestimate the sum of the extrapolated deaths while we still underestimate the overall number of 16 248 in the situation report.

While we see a slight decrease in reported cases towards the end of November, our model instead predicts a slight but further increase of infections; see Figure 13. This predicted increase is however consistent with the increasing number of confirmed cases in the beginning of December; cf. [73].

The slight decrease in confirmed cases end of November is likely to be a statistical artifact obtained due to a changed test strategy that has been in place in Germany since November 11 [79]. After that, the conducted PCR tests fell by around 200,000, while the positive rate rose from 7.86 percent to nine percent. Again, the curve of predicted intensive care patients lies above the extrapolated data. Our model thus shows the expected behaviour in times of higher infections.

#### 5.3.5 Summary of retrospective results

All in all, we have reached our aims: The median of our death and infection rates is close to the extrapolated real data without further adaptations for each individual scenario. We can identify the mean effectiveness of the sets of non-pharmaceutical interventions during different phases of the pandemic in Germany. Although we have no mechanism to predict local stochastic super-spreading events, we see how commuter activities lead to the spread of the disease along the routes visualized in Figure 2 (right). We can show how the commuter activities transfer the spread of local hot spots into their surrounding regions and we can predict regions with larger incidence over time spans of several weeks.

### 5.4 Predictive Scenario

We present twelve prospective scenarios for Germany from December 12 to January 10. In these scenarios, we assess different strictness and compliance levels in relation to the recently announced lockdown [16]. We assume that the strictness of the lockdown, the strictness of the partial lifting of the lockdown over Christmas, and the number of infected individuals (detected & undetected) will have the most influence on the effective mitigation and prediction of the spread of SARS-CoV-2 in that period.

Our aim is to predict the infected individuals in Germany from December 12 to January 10 while applying a strict lockdown from December 16 onward. We assume an effective contact reduction of this lockdown of 74 % or 67 %. From December 12 to December 16, we use the same contact reduction (50 %) as in our November scenario in Section 5.2. In our scenarios the lockdown restrictions are lifted for December 24-26 with contact reductions as given in Figure 15. Our predictions are based on an assumed initial number of undetected cases on December 12, namely 50 % and 150 %. Since the effects on the number of deaths through contact reduction in this period will only be seen from mid of January onward, we do not provide predictions for the number of deaths.

**Fig. 15.**
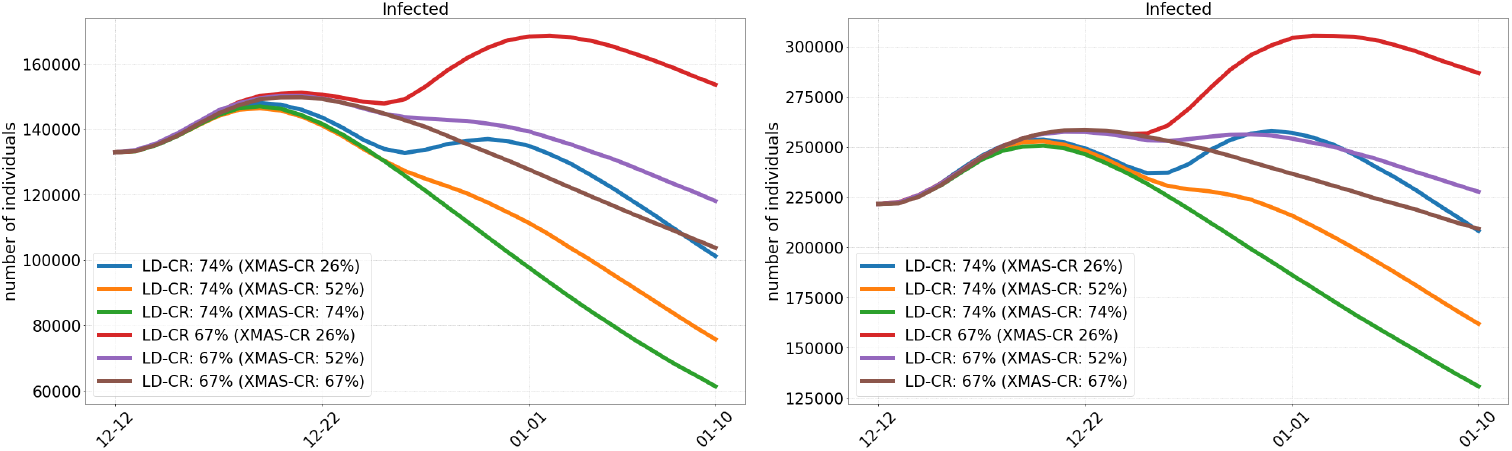
Prediction of the infected individuals in Germany from December 12 to January 10, applying a strict lockdown (LD) from December 16 and based on different potential numbers of undetected cases on December 12 (50 % left and 150 % right). Lifting of the lockdown restrictions for December 24-26 (XMAS) with lower contact reductions (CR) as presented.

### 5.5 Discussion of Predictive Scenarios

In Figure 15, we present scenarios considering contact reductions of 74 % and 67 % during lockdown with different increases in contact rates over Christmas. The scenarios on the left assume 50 % of undetected infections while the scenarios on the right assume 150 % of undetected infections on December 12.

Qualitatively, the curves in Figure 15 are similar for both proportions, 50 % (left) and 150 % (right), of undetected infections, but the number of infections differ considerably. We see that the strictest measures with 74 % contact reduction (CR) during lockdown and 74 % (green) or 52 % (orange) reduction over Christmas lead to the lowest number of infections by January 10. With a contact reduction of 67 % during the lockdown and over Christmas (brown), the number of infections also declines, but not to the same extent as in the green or orange scenario. The purple curve (52 % CR over Christmas) behaves to the brown scenario as the orange to the green scenario. If we only achieve 26 % contact reduction over Christmas (blue or red), there will be a substantial new rise of the number of infections before the lockdown can take its effect.

We observe that only slight additional contact reductions during lockdown can substantially change the speed of infection mitigation. Considering a lifting of restrictions over Christmas to allow for a mean of 7.2 daily contacts (CR 26 %) could throw back the containment of the virus for weeks. Note that these 7.2 daily contacts cannot be directly translated to persons in a room but highly depend on further parameters like (the type of) masks, distancing, ventilation etc. Considering the contact reduction of 52 % which may be closer to the NPIs in place over Christmas, we see that they also contribute to the spread the virus but far less than in the 26 % scenario.

## 6 Conclusion

We have extended the basic SECIR model [53] by age-resolution (Section 2.1) and added a model to resolve spatial heterogeneity (Section 2.2) using mobility data from the German Federal Employment Agency [10] and Twitter [85]. A numerical solution approach for the regional models has been presented in Section 2.3. One important future achievement will be the release of our modular epidemics simulation software *MEMILIO*, which is continuously under development [1] and uses the presented model and data. Our open-source software will also be available via a web front-end that can be used by the public and decision makers.

We have collected extensive data on epidemiological parameters and conducted own analyses based on patient-specific data; see Section 4. We have run experiments with different contact reduction factors and thus have identified parameters for the effectiveness of mitigation during different phases of the pandemic in Germany.

While age-resolved incidence data is a good starting point, further data such as daily number of tests per age group, age-resolved hospitalization and intensive care data would be valuable information to further validate our model.

In the retrospective scenarios the trends of infections have been reproduced well with the derived factors on the effective mitigation strategy in Germany. Local stochastic events such as super-spreading, which are naturally random, cannot be predicted. However, once infection information is included in the starting data, we see how commuter activities lead to increased infections also in surrounding counties. On the other hand, we see that less affected regions like the center of Germany or Northern and North-Eastern Germany are also predicted to be less affected by our simulations. Infection clusters as, e.g., appeared in large areas of Southern and Western Germany from beginning of autumn could be reproduced.

Further, we provide prospective scenarios (Fig. 15) for the current lockdown which differ in their strictness and compliance levels during lockdown as well as in the change of behavior over Christmas. For either 50 % or 150 % of undetected cases on December 12, we see some quantitative but no significant qualitative differences in the development of the scenarios. In the best case scenario, we expect a reduction of the infection numbers by a factor of up to two. With less severe restrictions (or less adherence to severe restrictions), this factor comes down to about 1.4 or 1.3. We see how increased contact rates (i.e. lower contact reduction numbers) over only three days of Christmas can substantially slow down the mitigation of the virus spread. If most of the restrictions are lifted over Christmas, virus containment will be set back by at least two weeks.

## Availability of data and material

The data used was mainly obtained from publicly available resources and references are provided. Data used in the software will be made publicly available with the software framework.

## Code availability

The software was developed by the authors and will be made publicly available open-source software in the near future.

## Acknowledgements

We thank Valerie Grappendorf, a student at the Hochschule für Gestaltung Schwäbisch Gmünd, for contributing Figure 3.

We express our deep gratitude to all study teams supporting the LEOSS study. The LEOSS study group contributed at least 5 per mille to the analyses of this study: Technical University of Munich (Christoph Spinner), University Hospital Freiburg (Siegbert Rieg), University Hospital Regensburg (Frank Hanses), Hospital Ingolstadt (Stefan Borgmann), Hospital Dortmund (Martin Hower), University Hospital Frankfurt (Maria Vehreschild), University Hospital Jena (Maria Madeleine Rüthrich), Hospital Ernst von Bergmann (Lukas Tometten), Hospital Bremen-Center (Christiane Piepel), University Hospital Essen (Sebastian Dolff), Johannes Wesling Hospital Minden (Kai Wille), Hospital Passau (Julia Lanzster), University Hospital Munich/ LMU (Michael von Bergwelt-Baildon), University Hospital Heidelberg (Uta Merle), University Hospital Augsburg (Christoph Rö mmele), Municipal Hospital Karlsruhe (Christian Degenhardt), University Hospital Erlangen (Julia Fürst), University Hospital Tübingen (Silvio Dalin), University Hospital Würzburg (Nora Isberner), University Hospital Ulm (Beate Grüner), University Hospital Cologne (Norma Jung), Cardiology and Intensive Care Medicine (Hendrik Haake), St. Josef-Hospital - Catholic Hospital Bochum (Kerstin Hellwig), Kreuzcher Diakonia (Wolfgang Rimili), Travel Clinic Paul-Lechler HospitalTübingen (Claudia Raichle), Hospital Leverkusen (Lukas Eberwein), Center for Infectiology Berlin/Prenzlauer Berg (Stephan Grunwald), Hacettepe University (Murat Akova), University Hospital Schleswig-Holstein - Kiel (Anette Friedrichs), Bundeswehr Hospital Koblenz (Dominic Rauschning), Practice at Ebertplatz (Christopf Wyen), University Hospital Düsseldorf (Bjö rn Jensen), University Hospital Dresden (Katja de With), Clinic Munich (Wolfgang Guggemos), Braunschweig Hospital (Jan Kielstein), Marien Hospital Herne, University Hospital Bochum (Beate Schultheis), Justus-Liebig-University Giessen (Jani Trauth), University Hospital Saarland (Robert Bals), Hospital Fulda (Philipp Markart), Petrus Hospital Wuppertal (Sven Stieglitz), Richmond Research Institute (Isobel Atkin), Maltese Hospital St. Franziskus-Hospital Flensburg (Mile Milovanovic), Robert-Bosch-Hospital Stuttgart (Katja Rothfuss), Sophien- and Hufeland Clinic Weimar (Jessica Rüddel), University Hospital Bonn (Jacob Nattermann), Agaplesion Diaconia Hospial Rotenburg (David Heigener), Hospital St. Joseph-Stift Dresden (Lorenz Walter), Elbland Hospital Riesa (Jö rg Schubert), St. Josef Hospital Kupferdreh (Joachim Voigt), Practice for general medicine Drs. Elisabeth Schrö dter & Gabriele Müller-Jö rger (Gabriele Müller-Jö rger), Helios-Hospital Pir (Christian Riedel), Oberlausitz-Hospital (Maximilian Worm). The LEOSS study infrastructure group: Jorg Janne Vehreschild (Goethe University Frankfurt), Lisa Pilgram (Goethe University Frankfurt), Melanie Stecher (University Hospital of Cologne), Max Schons (University Hospital of Cologne), Carolin E. M. Jakob (University Hospital of Cologne), Annika Claßen (University Hospital of Cologne), Susana M. Nunes de Miranda (University Hospital of Cologne), Sandra Fuhrmann (University Hospital of Cologne), Bernd Franke (University Hospital of Cologne), Nick Schulze (University Hospital of Cologne), Fabian Praßer (Charité, Universitätsmedizin Berlin) und Martin Lablans (University Medical Center Mannheim). The LEOSS study was supported by the German Center for Infection Research (DZIF) and the Willy Robert Pitzer Foundation.

