## Supplementary figures for "Assessment of effective mitigation and prediction of the spread of SARS-CoV-2 in Germany using demographic information and spatial resolution"

### Appendix

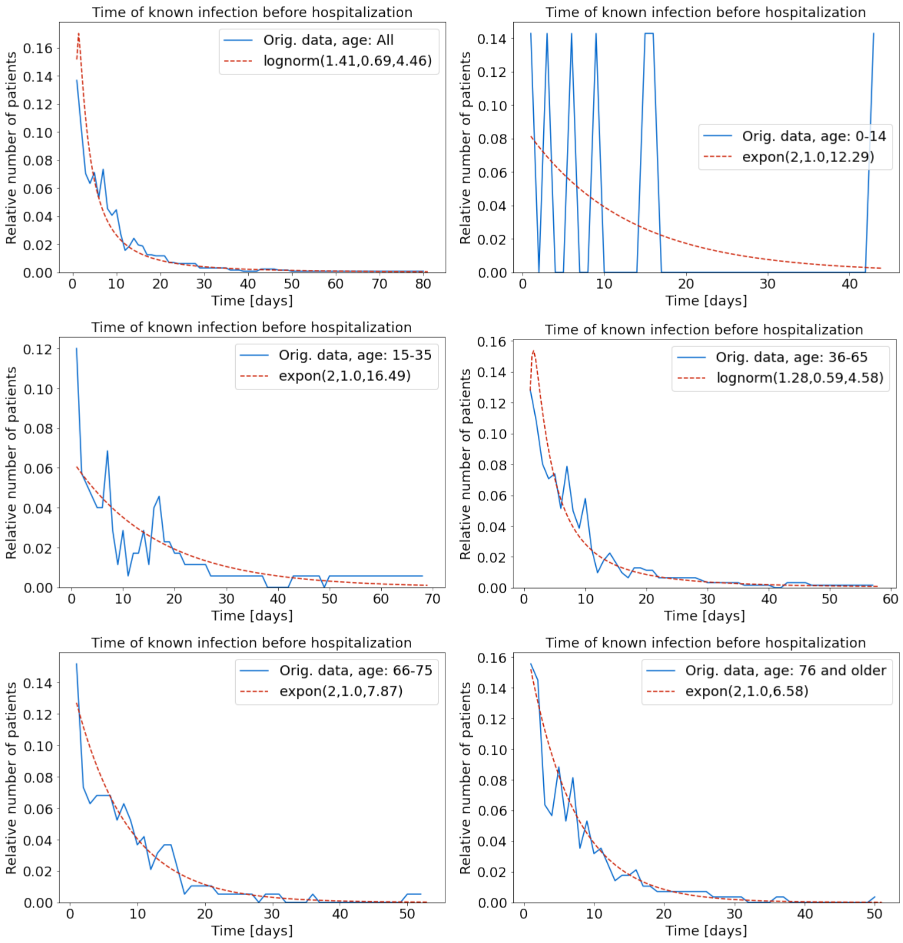


**Fig. 16** Graphs derived from LEOSS [58]. Age-specific time of known infection before admission to hospital. Best fit by either *lognormal distribution* or *exponential distribution* with given parameters.

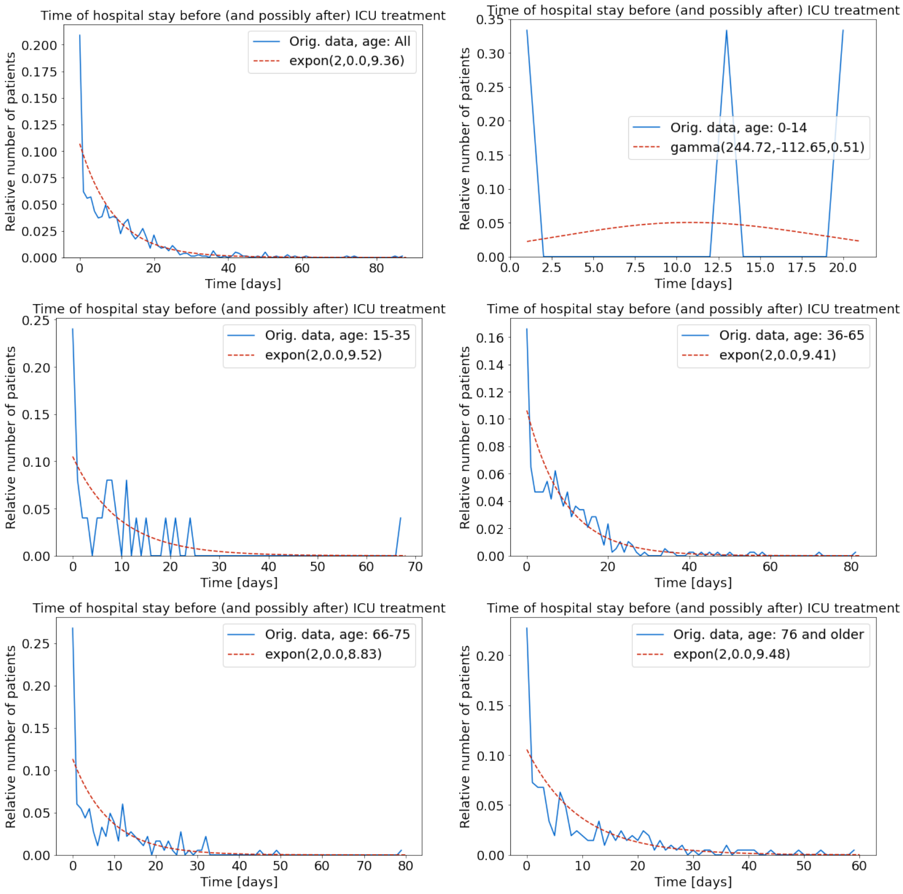


**Fig. 17** Graphs derived from LEOSS [58]. Age-specific time of hospitalization for patients needing intensiv care. It is however not clear how many days patients spent in hospital *before* and possibly *after* intensive care if eventually recovering from hospital. Best fit by *exponential distribution* with given parameters, for age group 0-14 years, gamma distribution was chosen but clearly lacking more data.

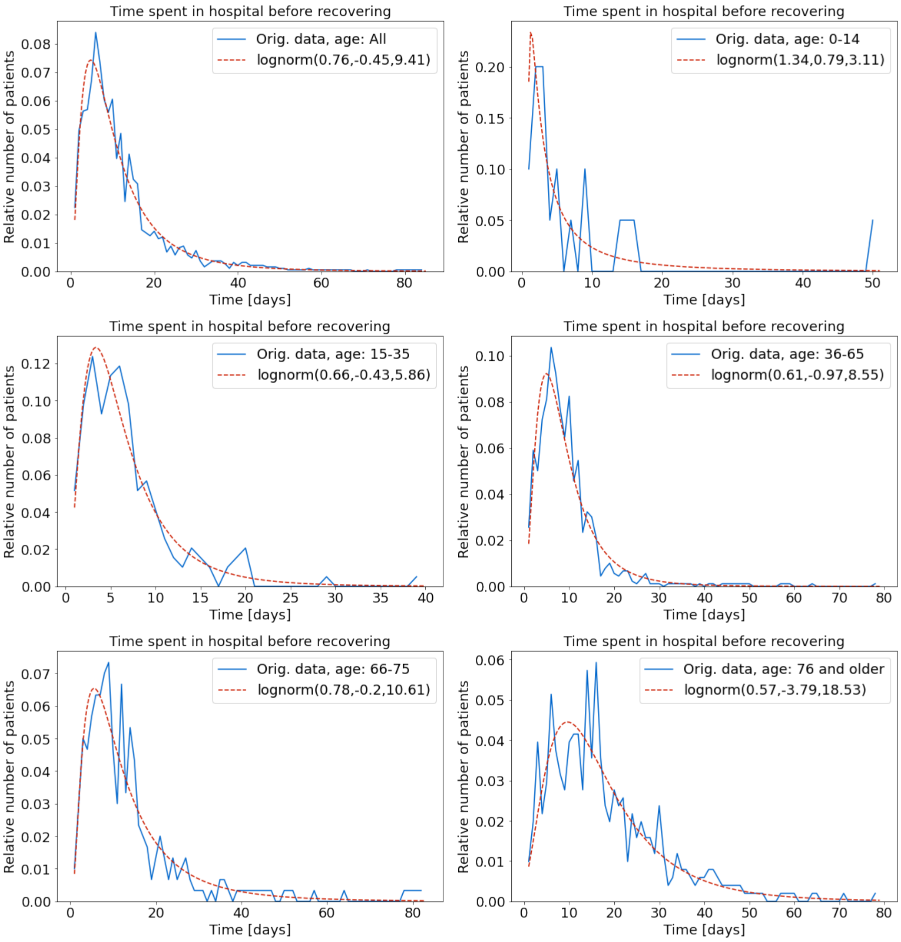


**Fig. 18** Graphs derived from LEOSS [58]. Age-specific time of hospitalization for patients not needing intensiv care. Best fit by *lognormal distribution* with given parameters.

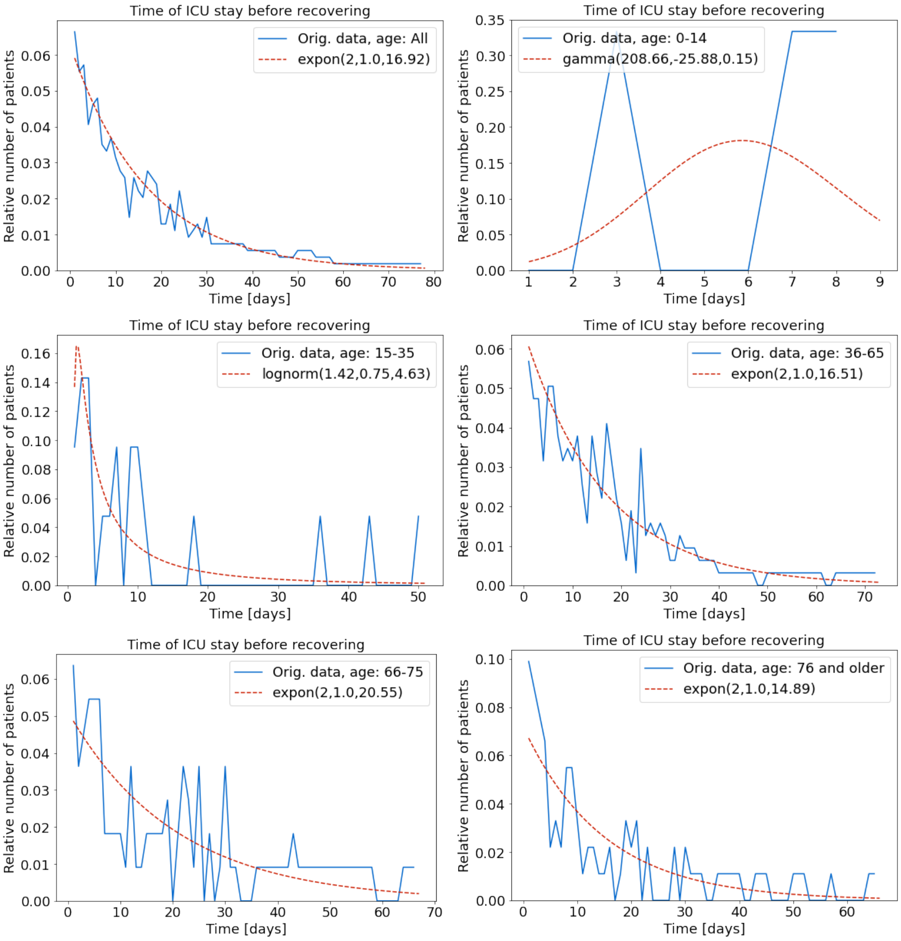


**Fig. 19** Graphs derived from LEOSS [58]. Age-specific time of intensive care before recovering. Best fit by either *lognormal distribution* or *exponential distribution* with given parameters. For age group 0-14 years, gamma distribution was chosen but clearly lacking more data.

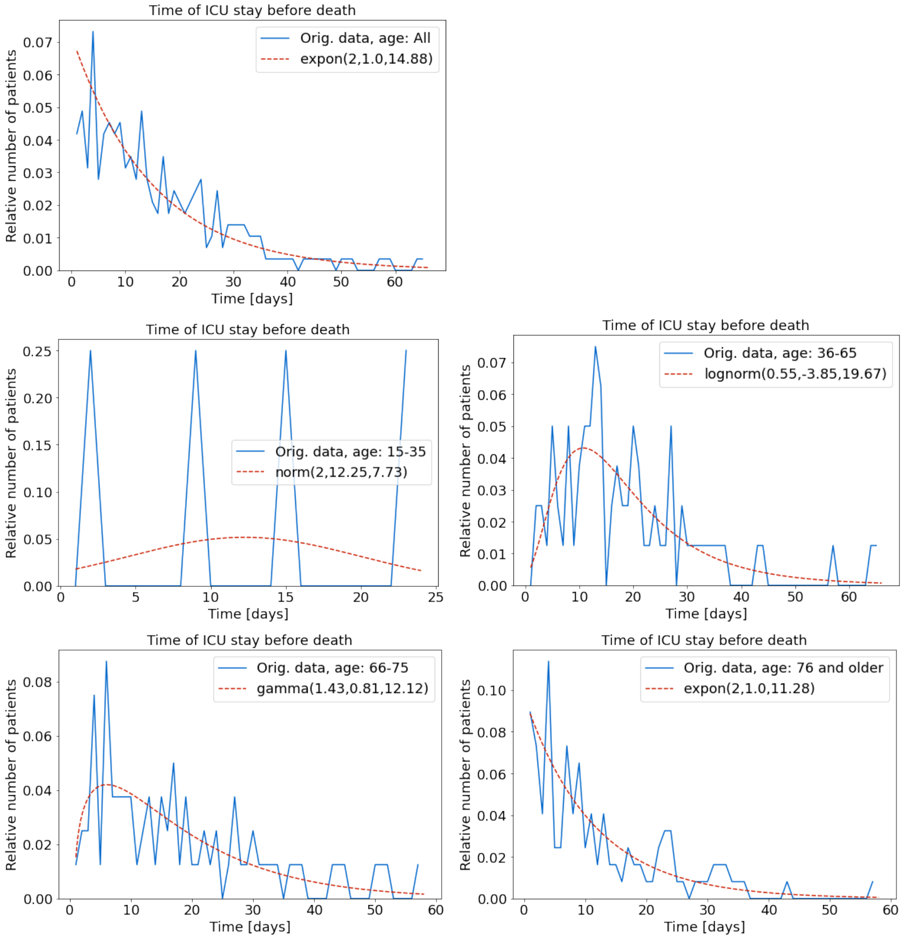


**Fig. 20** Graphs derived from LEOSS [58]. Age-specific time of intensive care before eventually dying. Best fit by either *exponential distribution*, *lognormal distribution* or *gamma distribution* with given parameters. For age group 0-14 years, no data is available, for age group 15-35, normal distribution was chosen but clearly lacking more data.

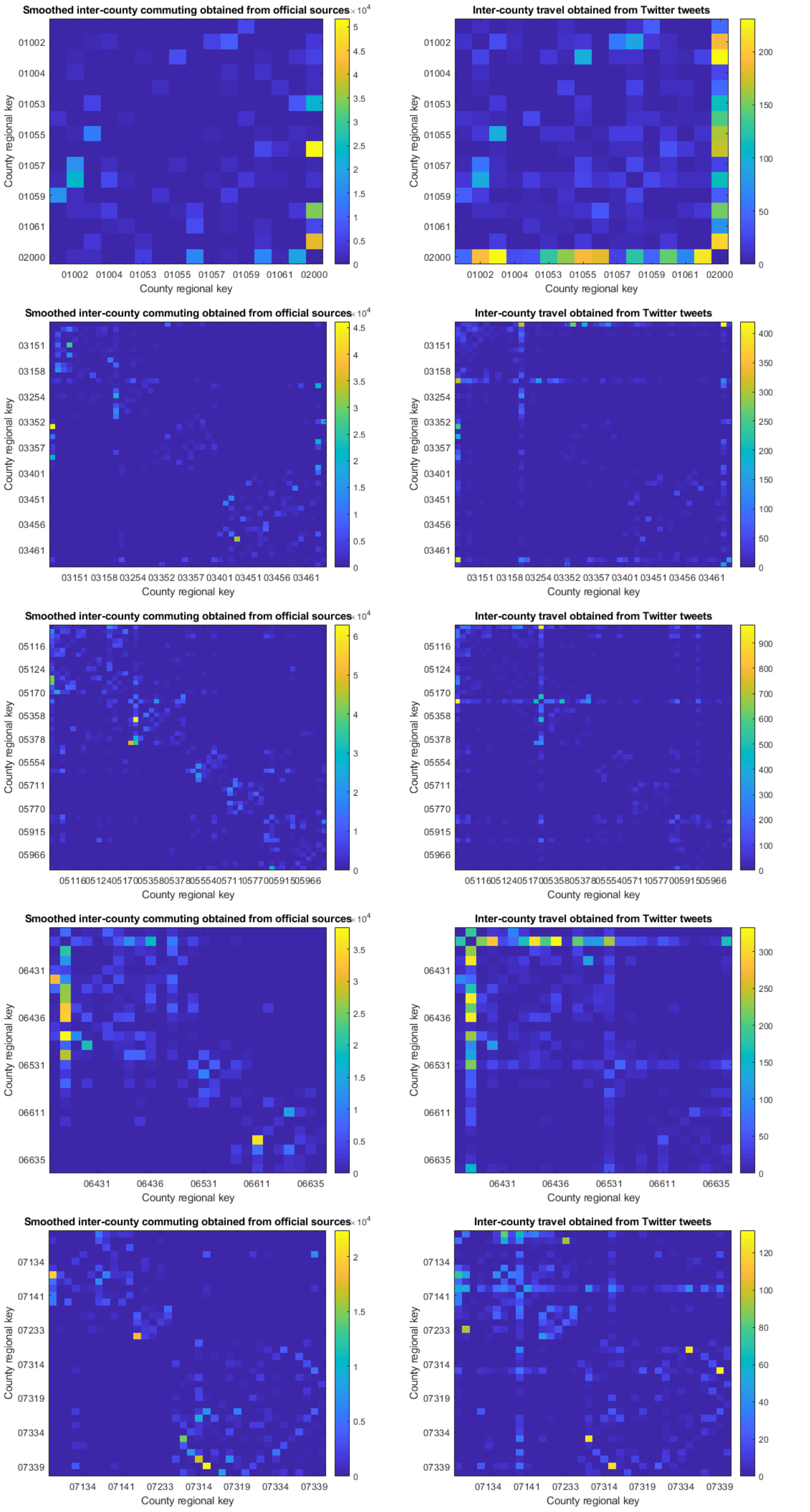


**Fig. 21** Mobility from [12] smoothed for long-distance commuting (left) and mobility obtained from geo-referenced tweets (right) as described in Section 2.2. Counties of different federal states and subgroups of federal states: Schleswig-Holstein to Rhineland-Palatinate.

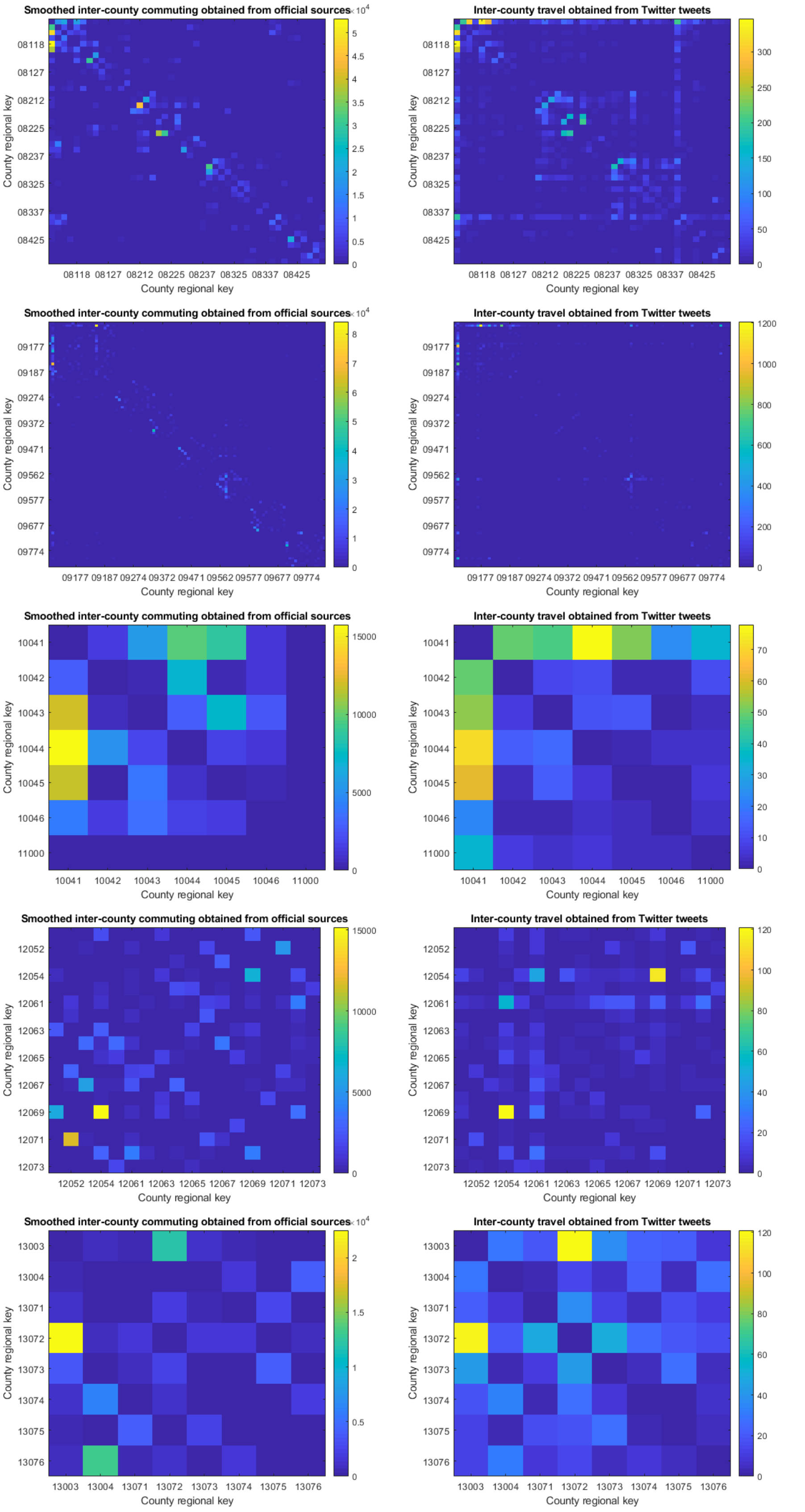


**Fig. 22** Mobility from [12] smoothed for long-distance commuting (left) and mobility obtained from geo-referenced tweets (right) as described in Section 2.2. Counties of different federal states: Baden-Württemberg to Mecklenburg-West Pomerania.

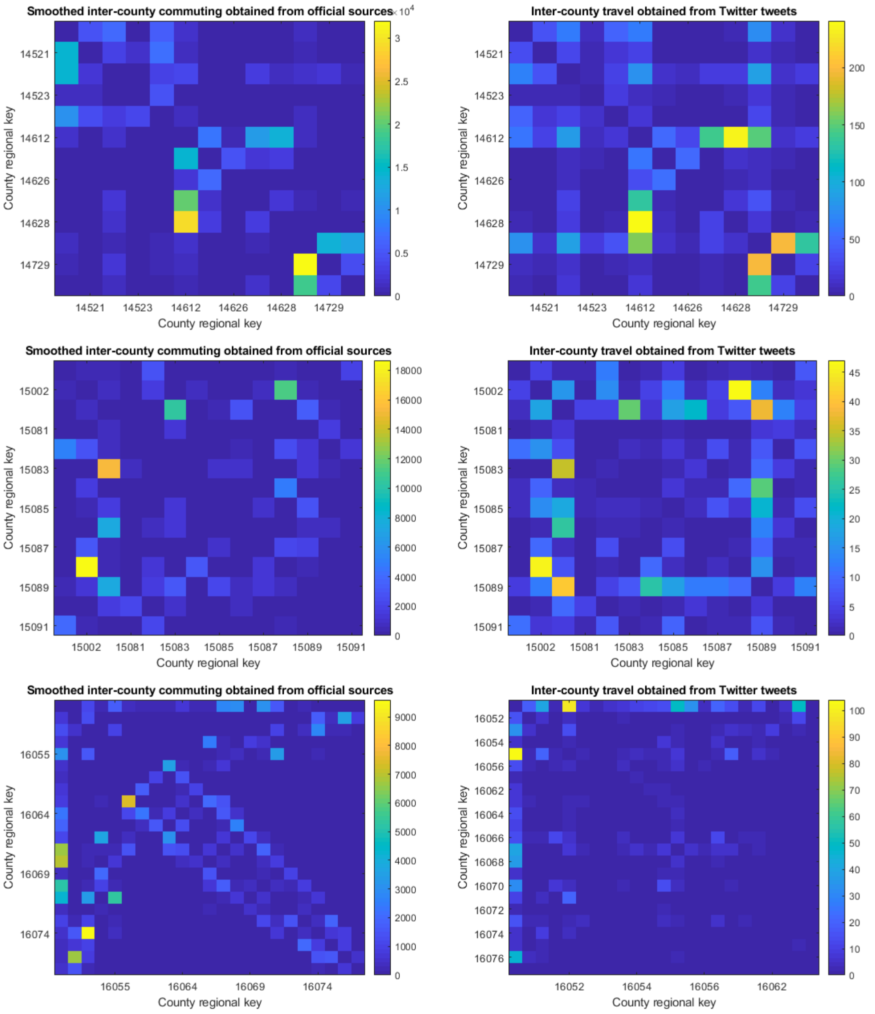


**Fig. 23** Mobility from [12] smoothed for long-distance commuting (left) and mobility obtained from geo-referenced tweets (right) as described in Section 2.2. Counties of different federal states: Saxony to Thuringia.
